# Charcot-Leyden Crystals drive pathology and inflammation in muco-obstructive lung disease

**DOI:** 10.1101/2025.11.26.25341000

**Authors:** Helena Aegerter, Iris Janssens, Kenneth Verstraete, Thomas Malfait, Ines Heyndrickx, Engi Ahmed, Jasmijn Burgelman, Hans Ulrichts, Tim Van Acker, Catelijne Stortelers, Siele Ceuppens, Sofie De Prijck, Manon Vanheerswynghels, Sam Dupont, Alysia Wayenberg, Ursula Smole, Femke Baeke, Amanda Goncalves, Tania Maes, Leander Jonckheere, Yannick Vande Weygaerde, Heidi Schaballie, Petra Schelstraete, Sebastian Riemann, Guy Brusselle, Richard Hoogenboom, Hue Vu, Annemieke Madder, Melia Magnen, Mark R. Looney, Wilfred Raymond, Annabelle Charbit, John V. Fahy, Savvas Savvides, Eva Van Braeckel, Bart N. Lambrecht

## Abstract

Mucus plugging leading to airway obstruction is a hallmark of chronic respiratory diseases, yet the biology of plugs and their contribution to airway inflammation remain poorly defined. Through guided retrieval and analysis of plugs from patients with allergic bronchopulmonary aspergillosis, we identify Charcot-Leyden crystals that are formed from eosinophil-derived Galectin-10 as ubiquitous and mechanistic components that generate highly localized type 2 inflammatory niches. These niches exhibit mixed granulocytic infiltration, including neutrophil activation and extracellular trap formation that reinforces mucus tenacity. In a mouse model recapitulating these characteristics, plugs containing Charcot-Leyden crystals induced persistent obstruction and inflammation, whereas systemic administration of a crystal-dissolving anti-Galectin-10 antibody markedly reduced crystal burden, inflammation and airway plugging. We further establish Galectin-10 as a sensitive biomarker of acute disease and develop a positron emission tomography approach enabling noninvasive visualization of crystal-rich plugs. These findings position Galectin-10 as a central driver and tractable therapeutic target in muco-obstructive lung disease.

## Introduction

Airway obstruction caused by mucus plugging plays a key role in the most common chronic respiratory diseases, such as asthma and chronic obstructive pulmonary disease (COPD)^1–3^. Despite their presumed prevalence in millions of people worldwide, mucus plugs have historically received limited attention, due to anatomical inaccessibility. Until recently, plugs could only be studied post-mortem or following invasive surgical or endoscopic procedures^4,5^. Advances in imaging now enable in situ quantification of mucus plugs, revealing their pervasiveness, even in mild disease^1,6^. High mucus plug burden correlates with airflow limitation (low forced expiratory volume (FEV₁) on lung function testing) and global markers of inflammation^1–3^, yet the local biological processes sustaining these obstructions remain largely uncharacterized. The chronic persistence of mucus plugs and their poor responsiveness to available treatments make their investigation an urgent clinical priority^1,2,7,8^.

Tenacious mucus plugging is especially prominent in Type-2-high, eosinophilic airway diseases such as asthma, chronic rhinosinusitis with nasal polyposis (CRSwNP), and allergic bronchopulmonary aspergillosis (ABPA). Activated eosinophils release the cytoplasmic protein Galectin-10, which spontaneously crystallizes to form Charcot-Leyden crystals (CLCs)^9^. These hexagonal bipyramidal protein crystals were first described over 150 years ago^10,11^. Although long regarded as inert byproducts of eosinophilic inflammation, recent studies suggest that CLCs can directly induce mucus production and promote inflammation^12^. However, the frequency and biological significance of CLCs within airway mucus plugs remain poorly defined. Technical limitations in accessing intact plugs, together with the species restriction of Galectin-10 expression to humans and select primates, have hindered direct mechanistic studies^13^.

ABPA is a co-morbidity of asthma or cystic fibrosis (CF) which presents with sensitisation to *Aspergillus* species^14–19^. This intensely eosinophilic disease presents with large, tenacious plugs, offering a unique clinical opportunity to study the biology of mucus plugs^14–16^. Previous reports have noted the presence of CLCs in ABPA samples, but their abundance, spatial localization, and functional impact have not been systematically characterized^4^.

Here, we combine analysis of human airway plugs in ABPA and a complementary mouse model to define the role of CLCs in mucus obstruction and inflammation. We show that, in contrast to nearby unplugged airways, airways with plugs containing CLCs represent sites of Type-2 high inflammation that are accompanied by intense granulocytic infiltration. We identify CLCs as a novel and sensitive biomarker in acute ABPA, that can be specifically dissolved to ameliorate local inflammation, and reduce mucus plugging. These findings identify CLC-containing mucus plugs as key inflammatory niches within the airway and establish Galectin-10 as a mechanistic driver and therapeutic target in muco-obstructive lung disease.

## Results

### Charcot-Leyden crystals and Galectin-10 as defining features of mucus plugs in ABPA

Twenty-three patients with acute allergic bronchopulmonary aspergillosis (ABPA) meeting the revised ISHAM-ABPA Working Group criteria were prospectively enrolled in a monocentric proof-of-concept study (Table 1, Supplementary Table 1). These patients were stratified based on their pre-disposing condition, asthma or CF, yet no significant differences were observed in immunological or serological profiles between the two underlying diseases (Table 2).

**Table 1.** Patient characteristics stratified by study group. Abbreviations: ABPA: allergic bronchopulmonary aspergillosis; Af: Aspergillus fumigatus; CF: cystic fibrosis; CFTR: cystic fibrosis transmembrane conductance regulator protein; ELX: elexacaftor; FeNO: fractional exhaled nitric oxide; FEV1 %pred: percent of predicted of forced expiratory volume in one second, FVC %pred: percent of predicted of forced vital capacity; ICS: inhaled corticosteroids; Ig: immunoglobulin; OCS: oral corticosteroids; ppb: parts per billion; WBC: white blood cell count.° Indication for oral or intravenous antibiotics. °°benralizumab (n=2), mepolizumab (n=2), dupilumab (n=2).°°°dupilumab (n=1), tezepelumab (n=1). *Cut-off value Af IgE 0.35 kU/L. **If a post bronchodilator value was not available, the pre bronchodilator value was used. ***All subjects without cystic fibrosis have asthma. Statistical test for continuous variables: Mann-Whitney U test; for categorical variables: Fisher’s Exact test. P values < 0.05 are in bold. Note1: five individuals were included in both groups, and one individual was included once in ‘no acute ABPA’ and twice in ‘acute ABPA’. Each entry represents a unique clinical episode, not a unique individual. Note2: eight individuals provided a sputum and a plug sample within the same clinical episode, of whom four in ‘no acute ABPA’ and four in ‘acute ABPA’.

| Study group<br>n | no acute ABPA<br>132 | acute ABPA<br>23 | p value |
| --- | --- | --- | --- |
| Age (years), mean (SD) | 33 (15) | 43 (22) | **0.044** |
| Child 2-12 years, n (%) | 4 (3.0) | 3 (13.0) | 0.067 |
| Female sex, n (%) | 66 (50.0) | 10 (43.5) | 0.654 |
| Post-bronchodilator spirometry** |  |  |  |
| FEV<sub>1</sub>, %pred, mean (SD) | 80.6 (21.6) | 78.7 (17.7) | 0.704 |
| FVC, %pred, mean (SD) | 93.4 (15.3) | 95.4 (12.5) | 0.434 |
| FEV<sub>1</sub>/FVC, %, mean (SD) | 70.2 (13.7) | 65.4 (9.9) | **0.049** |
| CF***, n (%) | 110 (83.3) | 9 (39.3) | **<0.001** |
| CF, pulmonary exacerbation current°, n (%) | 12 (10.9) | 0 (0.0) | 1 |
| CFTR modulation, n (%) | 83 (75.5) | 4 (44.4) | 0.058 |
| History of ABPA, n (%) | 35 (26.5) | 16 (69.6) | **<0.001** |
| Sensitization for Af* | 76 (57.6) | 23 (100.0) | **<0.001** |
| Blood eosinophils, x10<sup>6</sup> cells/μL, mean (SD) | 261 (281) | 802 (674) | **<0.001** |
| Spontaneous sputum eosinophils, % | 2.8 (7.1) | 50.6 (45.0) | **0.002** |
| FeNO, ppb | 20.7 (24.3) | 40.8 (32.6) | **<0.001** |
| Total IgE (kU/L), mean (SD) | 412.6 (1575.0) | 3047.9 (4196.8) | **<0.001** |
| Af IgE (kU/L), mean (SD) | 5.8 (13.4) | 22.6 (20.1) | **<0.001** |
| Af IgG (mg/L), mean (SD) | 43.2 (39.8) | 197.4 (155.2) | **<0.001** |
| ICS maintenance, n (%) | 83 (62.9) | 19 (82.6) | 0.094 |
| OCS use, n (%) | 1 (0.8) | 3 (13.0) | **0.011** |
| Anti-T2 biologic, n (%) |  |  |  |
| omalizumab | 11 (8.3) | 3 (13.0) | 0.439 |
| other | 6<sup>oo</sup> (4.5) | 2<sup>ooo</sup> (8.6) | NA |
| Number of study samples, n (%) |  |  |  |
| Serum | 130 (98.5) | 22 (95.7) | 0.384 |
| Spontaneous sputum | 45 (34.1) | 5 (21.7) | 0.335 |
| Bronchoscopic mucus plug | 9 (6.8) | 15 (65.2) | **<0.001** |

**Table 2.** Patient characteristics of acute ABPA patients, stratified by underlying disease. Abbreviations: ABPA: allergic bronchopulmonary aspergillosis; *Af*: *Aspergillus fumigatus*; CF: cystic fibrosis; FeNO: fractional exhaled nitric oxide; FEV1 *%pred*: percent of predicted of forced expiratory volume in one second; ppb: parts per billion;*Cut-off value Af IgE 0.35 kU/L. **If a post bronchodilator value was not available, the pre bronchodilator value was used. Statistical test for continuous variables: Mann-Whitney U test; for categorical variables: Fisher’s Exact test. P values < 0.05 are in bold. Note1: two individuals were included twice.. Each entry represents a unique clinical episode, not a unique individual. Note2: four individuals provided a sputum and a plug sample within the

| Study group<br>Underlying condition<br>n (group %) | acute ABPA |  |  |  |
| --- | --- | --- | --- | --- |
|  | Total<br>23 | CF<br>9 (39.1) | asthma<br>14 (60.9) | p value |
| Age (years), mean (SD) | 43 (22) | 19 (9) | 58 (12) | **<0.001** |
| Child 2-12 years, n (%) | 3 (13.0) | 3 (33.3) | 0 (0.0) | **0.047** |
| Female sex, n (%) | 10 (43.5) | 1 (11.1) | 9 (64.3) | **0.029** |
| Post-bronchodilator spirometry** |  |  |  |  |
| FEV<sub>1</sub>, %pred, mean (SD) | 78.7 (17.7) | 68.1 (21.2) | 86.1 (10.4) | **0.041** |
| FVC, %pred, mean (SD) | 95.4 (12.5) | 89.3 (14.7) | 99.5 (9.1) | 0.116 |
| FEV<sub>1</sub>/FVC, %, mean (SD) | 65.4 (9.9) | 63.8 (13.7) | 66.5 (6.5) | 0.973 |
| History of ABPA, n (%) | 16 (69.6) | 7 (77.8) | 9 (64.3) | 0.657 |
| Sensitization for *Af* * | 23 (100.0) | 9 (100.0) | 14 (100.0) | 1 |
| Blood eosinophils, x10<sup>6</sup> cells/μL, mean (SD) | 802 (674) | 855 (374) | 767 (825) | 0.108 |
| Spontaneous sputum eosinophils, % | 50.6 (45.0) | 2.8 (0.3) | 82.4 (15.4) | 0.083 |
| FeNO, ppb | 40.8 (32.6) | 43.9 (31.6) | 3.8 (34.4) | 0.787 |
| Total IgE (kU/L), mean (SD) | 3047.9 (4196.8) | 1667.0 (779.5) | 4003.8 (5291.3) | 0.616 |
| *Af* IgE (kU/L), mean (SD) | 22.6 (20.1) | 27.8 (14.2) | 18.9 (23.2) | 0.053 |
| *Af* IgG (mg/L), mean (SD) | 197.4 (155.2) | 232.3 (155.4) | 176.0 (157.3) | 0.169 |
| Number of study samples, n (%) |  |  |  |  |
| Serum | 22 (95.7) | 9 (100.0) | 13 (92.9) | 1 |
| Spontaneous sputum | 5 (21.7) | 2 (22.2) | 3 (21.4) | 1 |
| Bronchoscopic mucus plug | 15 (65.2) | 7 (77.8) | 8 (57.1) | 0.400 |

To introduce typical findings, a case study is presented in entirety: a 75-year old male with the diagnosis of acute ABPA as a comorbidity of severe asthma. High resolution computed tomography (HRCT) revealed extensive mucus plugging and central bronchiectasis typical of disease (Fig. 1A). Automated airway reconstructions in this patient showed, in this case, complete obliteration of the right lower main bronchus with total exclusion of ventilation yet no atelectasis ensued. Guided retrieval of mucus plugs identified by CT-scan was performed via bronchoscopy. Multiple plugs varying between 1 – 8 cm in length could be extracted from this single patient (Fig. 1B). These plugs consisted of hard and tenacious material which required mechanical disruption with scissors to process (Fig. 1B). Histological examination showed intense inflammation within and around the plug, with numerous hexagonal bipyramidal crystals up to ∼80 µm (Fig. 1C, Supplementary Fig. 1A, B). Immunofluorescence confirmed these structures as Galectin-10/Charcot-Leyden crystals (CLCs) (Fig. 1D). Scanning and transmission electron microscopy demonstrated CLCs integrated into the mucus network, with mucin strands intimately associated with crystal surfaces (Fig. 1E, Supplementary Fig. 1B).

**Figure 1:**
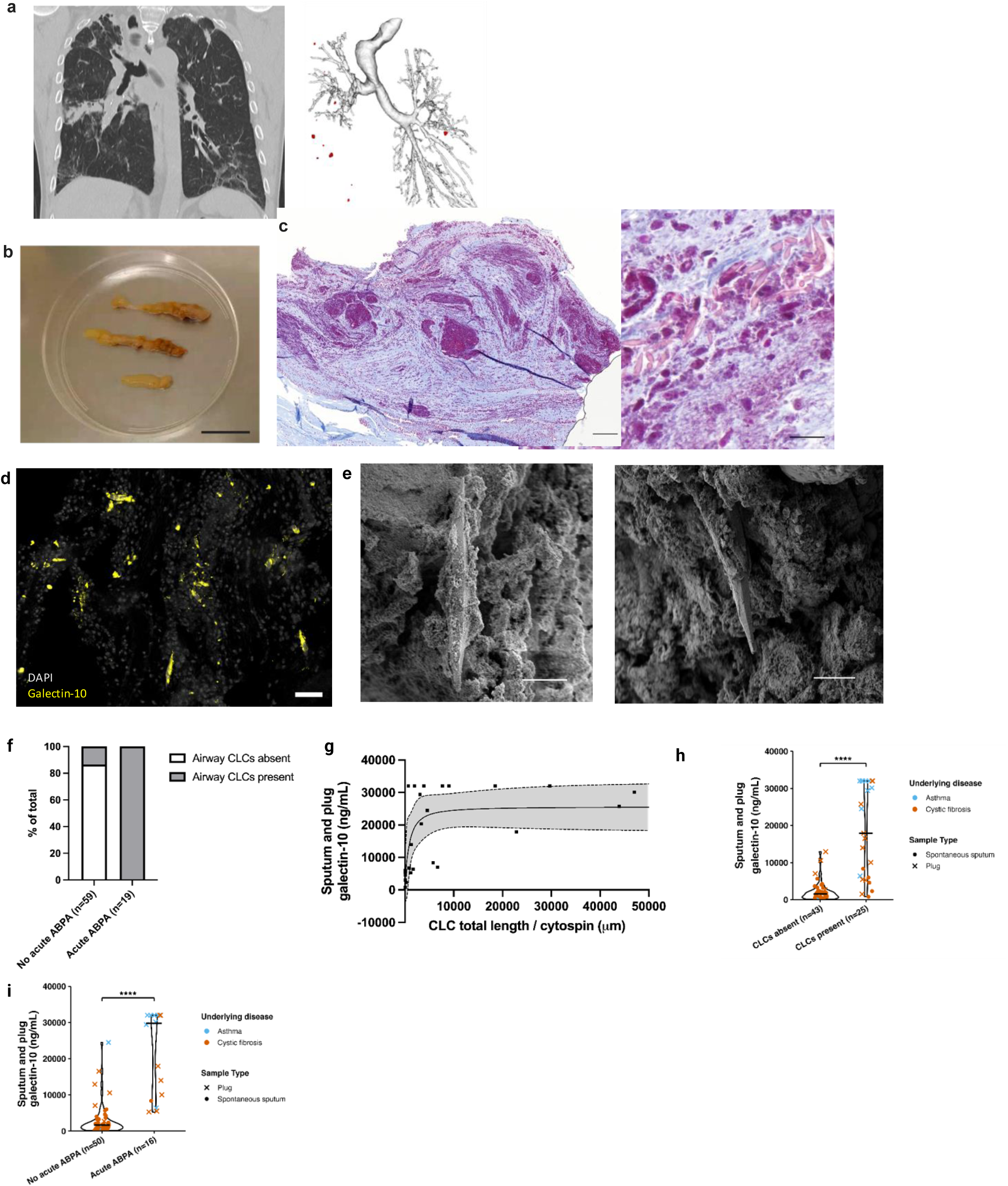
Charcot–Leyden crystals and Galectin-10 as defining features of mucus plugs in ABPA. **a**, Thorax CT scan reveals extensive mucus plugging in the right lower lobe of an ABPA patient (*left*). Automated rendering of the bronchial tree reveals total exclusion of the right lower lobe from ventilation (*right*). **b,** Macroscopic image of exemplar (but not all) mucus plugs extracted from the obliterated airways in panel a. Scale bars, 20 mm. **c,** Light microscopy image of a Masson’s trichrome staining of a mucus plug extracted from the airways of an ABPA patient. Charcot-Leyden crystals (CLCs) appear pale pink. Scale bars, 500 μm (overview image) and 100 μm (detail). **d,** Immune fluorescent staining of a mucus plug for galectin-10 (Gal-10) (yellow) and nuclei (DAPI, light grey). Scale bars, 20 μm. **e,** CLCs embedded in a sputum plug, image acquired by scanning electron microscopy. Scale bars, 10 μm. **f,** CLC presence in spontaneous sputum, mucus plugs or broncho-alveolar lavage fluid (one sample type shown per patient) as a sensi tive diagnostic marker for ABPA. **g,** Association between CLC burden and galectin-10 concentration in sputum. Dot plot showing CLC total length per cytospin versus sputum Gal-10 (ng mL⁻¹). A sigmoidal four-parameter logistic fit (solid line) indicates a positive, saturating relationship (R² = 0.47). Spearman correlation analysis shows a significant ass ociation (ρ = 0. 68, 95% CI 0.41–0.85, *p* < 0. 001). Each point represents an individual sample. Shaded area, 95% CI of the fit. **(g-i)** Higher limit of quantification for the Galectin-10 ELISA was 32,000ng mL⁻¹. **h,** Sputum and plug Gal-10 concentrat ion by CLC presence. Violin plots show Gal-10 in subjects without (n = 43, median 1,577 ng mL⁻¹) or with (n = 25, median 17,913 ng mL⁻¹) CLCs. *p* < 0. 0001. **i,** Sputum and plug Gal-10 concentration by acut e ABPA diagnosis. Violin plots show Gal-10 in subjects without (n = 50, median 1,679 ng mL⁻¹) or with (n = 16, median 29,769 ng mL⁻¹) acute ABPA. *p* < 0. 0001. **j,** Receiver operating characteristic (ROC) curves comparing investigational biomarkers (Gal –10 in sputum and plug, sputum alone and serum) and canonical biomarkers for acute ABPA diagnosis. Area under the curve (AUC) values are shown; all comparisons were statistically significant (*p* ≤ 0.001). ABPA, allergic bronchopulmonary aspergillosis; CLCs, Charcot-Leyden cryst als; CT, comput ed tomography; Gal-10, galectin-10. Points are colored by disease (asthma, blue; CF, orange) and shaped by sample type (spont aneous sput um, ●; plug, ×). Black crossbars indicate medians. Each sample represents one unique clinical episode. I f spont aneous sputum and plug from the same episode were available (n=8), the plug sample was selected. * *p* < 0. 05, ** *p* < 0. 01, *** *p* < 0. 001, **** *p* < 0. 0001. Mann-Whitney U statistical test unless otherwise stated. See Supplementary Table 1 for full statistical details in the different

The ubiquitous presence of CLCs within the plugs of acute ABPA patients prompted a more in depth analysis of prevalence within the entire group, taking advantage of all available sample types. In patients with acute ABPA, CLCs were present in at least one of the airway samples obtained (*i.e.* spontaneous sputum, mucus plug and/or broncho-alveolar lavage (BAL) fluid) (Fig. 1F, Supplementary Fig. 1C). This corresponds with a perfect sensitivity (1, 95% CI 0.82-1.00) and high specificity (0.86, 95% CI 0.75-0.94) for acute ABPA. Quantification of Galectin-10 levels across a singular airway sample per patient (spontaneous sputum or plug) demonstrated a strong correlation between soluble Galectin-10 concentration and total CLC length (manually quantified by cytospin), validating this ELISA for objective measurement of CLC burden (Fig. 1G). Samples containing CLCs exhibited markedly higher Galectin-10 than those without (Fig. 1H), and this proved to be a distinguishing feature of patients with acute ABPA (Fig. 1I, Supplementary Fig. 1D).

Together, these findings identify CLCs and Galectin-10 as abundant, quantifiable constituents of mucus plugs and sensitive biomarkers of acute ABPA.

### CLC-containing mucus plugs act as self-amplifying foci of type-2 inflammation

Obstructive airway diseases like ABPA, asthma, COPD and CF are usually considered as inflammatory diseases that affect the entire lung, yet mucus plugging can be seen to occur in discrete areas along the bronchial tree. We therefore hypothesized that mucus plugs might constitute hot spots for local inflammation. Biopsies were taken from both a plugged and unplugged airway of an individual patient to compare inflammation at these sites. Marked local eosinophil infiltration was observed exclusively around CLC-containing plugs (Fig. 2A-D), and occurred despite normal peripheral blood eosinophil counts in some patients (Supplementary Table 2, Supplementary Fig. 2A). This enhanced inflammatory niche around plugged airways was absent when the plugs lacked CLCs (Fig. 2D). These data support the idea that the presence of CLCs is the main driver of a local inflammatory niche around plugged airways.

**Figure 2:**
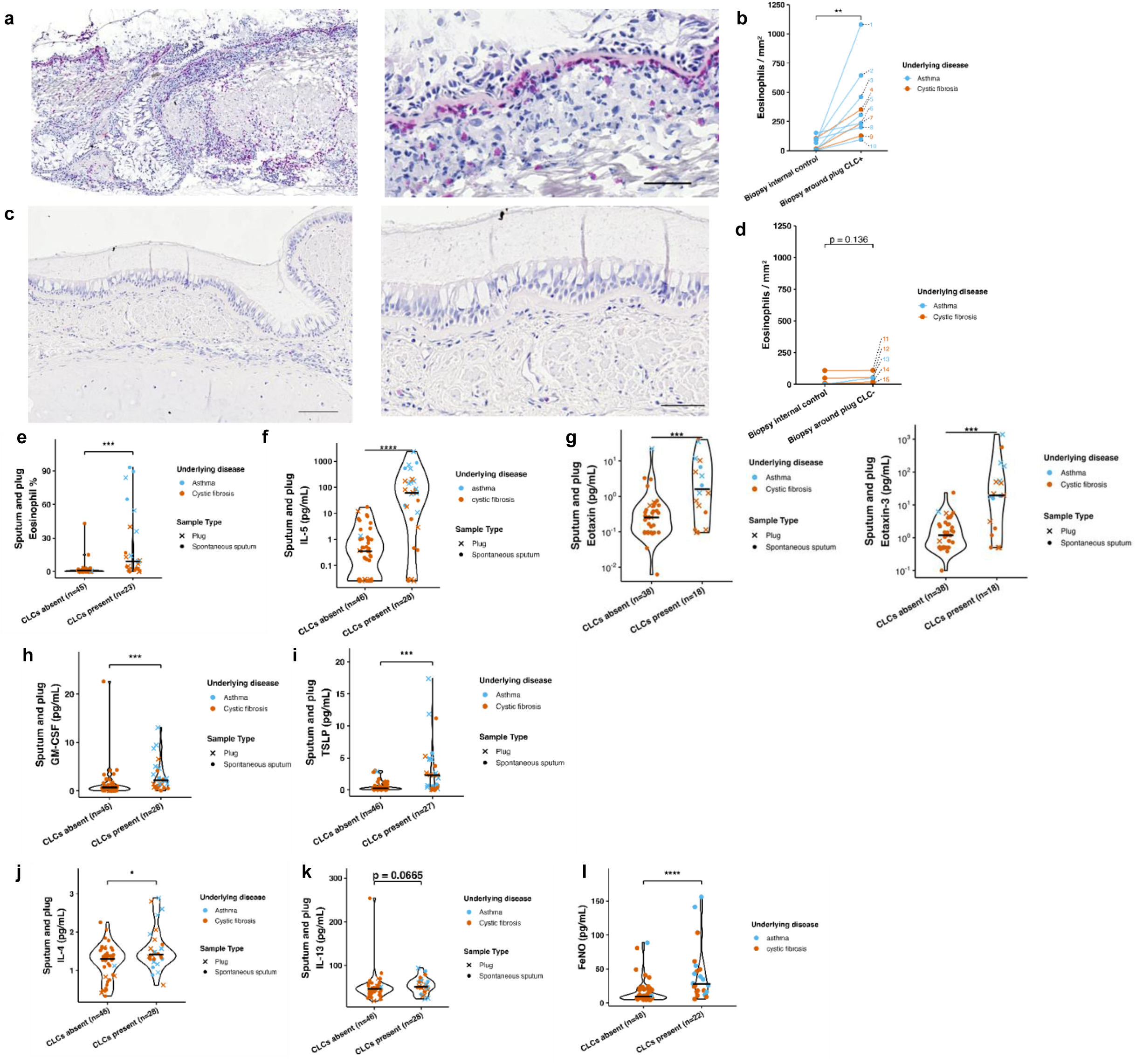
CLC-containing mucus plugs act as self-amplifying foci of type-2 inflammation. **a**, Light microscopic image of an H&E-stained bronchial mucosal biopsy taken from the surrounding of a mucus plug containing Charcot–Leyden cryst als (CLCs). Scale bars, 500 μm (overview), 20 μm (detail). **b,** Paired eosinophil counts in bronchial mucosal biopsies near CLC positive plugs and paired internal cont rols (median 271, eosi nophils/mm2) compared to bronchial mucosa from an internal unplugged control segment (median 43.3 eosinophils/mm2). Wilcoxon matched-pairs signed-rank test. *p* = 0. 002. **c,** Light microscopic image of an H&E-stained bronchial mucosal biopsy obtained in a contralateral internal unplugged cont rol segme nt. Scale bar, 500 μm (overview), 20 μm (detail). **d,** Paired eosinophil counts in bronchial mucosal biopsies near CLC negative plugs and paired internal controls (median 51 eosino phils/mm2) compared to bronchial mucosa from an internal unplugged control segment (median 8 eosinophils/mm2). Wilcoxon matched-pairs signed-rank test. *p* = 0. 136. Comparison of samples with CLCs absent or present in the context of asthma and CF for (**e**), (**f**), sputum and plug eosinophil percentage (median 0.72 vs 9.16%, *p* < 0. 0001); (**g**), (**h**), sputum and plug IL-5 (median 0.36 vs 61.59 pg mL⁻¹, *p* < 0. 0001) on a logarithmic scale; (**i**), sputum and plug eotaxin-1 and eotaxin-3; (**j**), sputum and plug GM-CSF (median 0.71 vs 2.18 pg mL⁻¹, *p* < 0. 001); (**k**), sputum and plug TSLP (median 0.26 vs 2.28 pg mL⁻¹, *p* < 0. 001); (**l**), sputum and plug IL-4 (median 1,30 vs 1,41 pg mL⁻¹, *p* < 0. 05); (**m**), sputum and plug IL-13 (median 46,16 vs 50.89 pg mL⁻¹, *p* < 0. 0001); (**n**), fractional exhaled nitric oxide (FeNO) (median 9.5-27.5 ppb, *p* < 0. 0001). ABPA, allergic bronchopulmonary aspergillosis; CLCs, Charcot-Leyden cryst als; Gal-10, galectin-10. Ppb, parts per billion. Points are colored by disease (asthma, sky blue; CF, vermilion) and shaped by sample type (spontaneous sputum,●; plug, ×). Black crossbars indicate medians. * *p* < 0. 05, ** *p* < 0. 01, *** *p* < 0. 001, **** *p* < 0. 0001. Mann-Whitney U statistical test unless otherwise stated.

Patients with CLCs in the airways also displayed airway eosinophilia, elevated concentrations of the eosinophil growth factors interleukin (IL)-5 and granulocyte-macrophage colony-stimulating factor (GM-CSF) and of eosinophil-recruitment chemokines, including eotaxin-1 and eotaxin-3 (Fig. 2E-H). The epithelial-derived cytokine thymic stromal lymphopoietin (TSLP), known to drive type 2 airway inflammation by activating dendritic cells (DCs) and innate lymphoid cells type 2 (ILC2s)^20^, was enriched in samples from CLC-rich plugged airways (Fig. 2I). A prominent type-2 cytokine signature was evident, characterized by increased IL-4, IL-13, and fractional exhaled nitric oxide (FeNO) (Fig. 2J-L), while type-1 cytokines tumor necrosis factor (TNF)-α and IL-1β were reduced (Fig. Supplementary Fig. 2B-C). With the exception of IL-5 (Supplementary Fig. 1D), this signature was confined to the airway, demonstrating that plugs containing CLCs establish localized inflammatory niches not mirrored systemically.

### CLC-induced neutrophil activation and NET formation reinforce mucus tenacity

Despite a dominant type-2 inflammatory milieu, neutrophilic infiltration was observed surrounding the CLC-containing plugs, accompanied by increased levels of the neutrophil-attracting chemokine IP-10. (Fig. 3A-B). Within the plug itself, intense neutrophil activation was observed in the vicinity of CLCs, with a diffuse myeloperoxidase (MPO)-DAPI co-staining indicative of the presence of Neutrophil extracellular traps (NETs) (Fig. 3C). Detection of NETs by ELISA demonstrated that CLC presence was associated with significantly increased NET production within plugs (Fig. 3D). NETs were more evident in mucus plugs than in spontaneous sputum (Supplementary Fig. 3A), and peripheral blood neutrophils were not elevated (data not shown).

**Figure 3:**
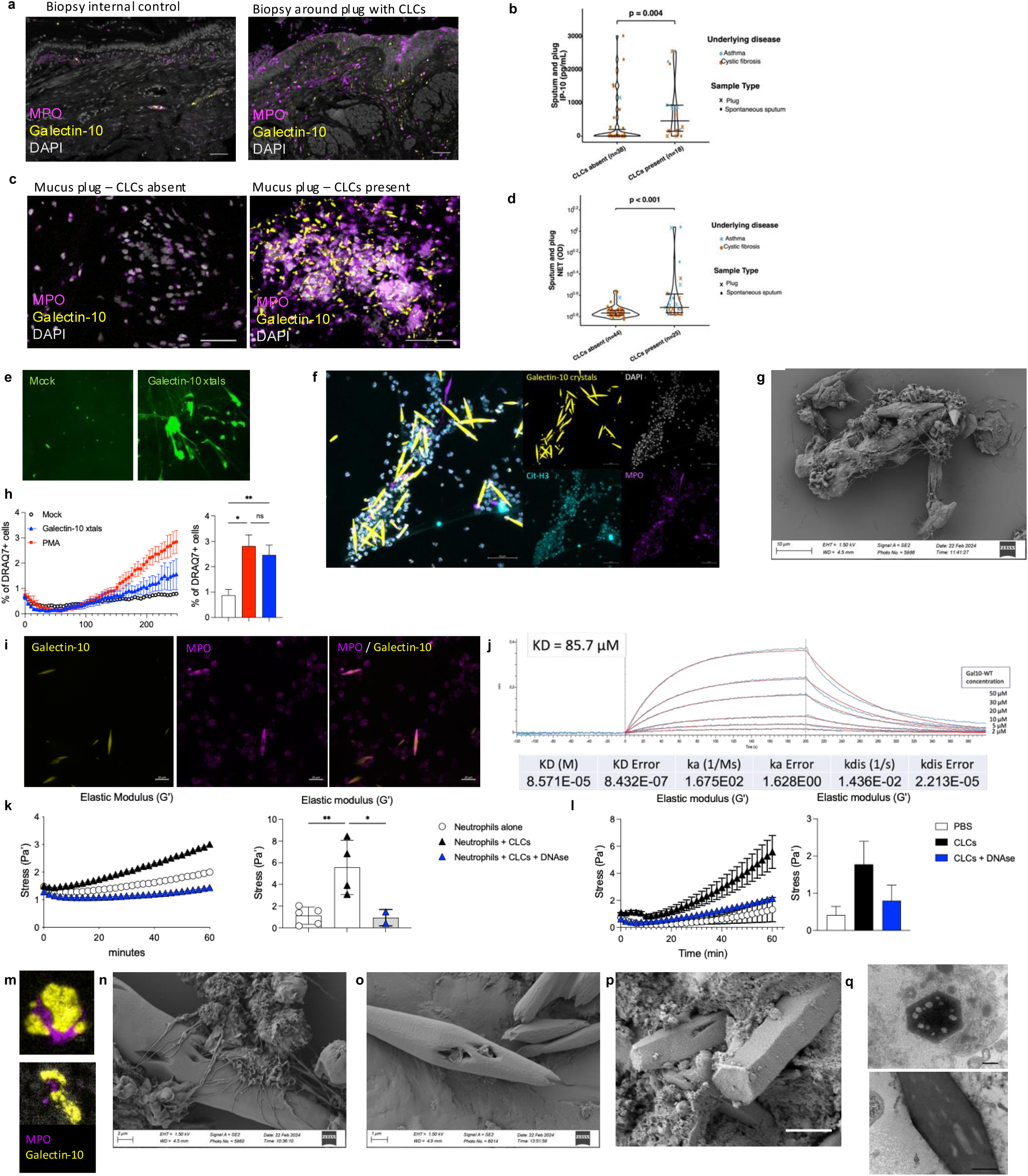
CLC-induced neutrophil activation and NET formation reinforce mucus tenacity. **a**, Immunostaining for neutrophils (MPO) in a mucosal biopsy taken from an internal control segment (left) and a plugged segment with CLCs present (right). **b,** Comparison of samples with CLCs absent or present in the cont ext of asthma and CF for IP-10 **c,** Immunostaining for Neutrophil Extracellular Traps (NETs) in mucus plugs without (left) and with (right) CLCs. **d,** Comparison of samples with CLCs absent or present in the context of asthma and CF for NETosis via ELISA (median 0.17 vs 0.19, *p* < 0. 0001). Points are colored by disease (asthma, sky blue; CF, vermilion) and shaped by sample type (spontaneous sputum, ●; plug, ×). Black crossbars indicate medians. Mann-Whitney U statistical test. **e,** Release of NETs by human peripheral neutrophils left unstimulated (PBS) or stimulated with 100µg/ml CLCs for 4 hours followed by staining of extracellular DNA with SYTOX Green Scale bar **f,** Detection of NETs produced by human neutrophils stimulated with 100µg/ml CLCs for 4 hours followed by immuno-staining of Citrullinated H3 (Cit-H3), Myeloperoxidase (MPO), DAPI and detection of FITC-CLCs Scale bar, 50µm **g,** Multiple neutrophils and NETs decorate a CLC following incubation of human neutrophils with 100µg/ml CLCs for 4 hours, shown with scanning electron microscopy scale bar **h,** Left panel demonstrates an example of production of extracellular DNA (DRAQ7) quantified by live cell imaging following incub ation of human peripheral neutrophils with medium (mock), PMA (100nm) or 100µg/ml CLCs for 4 hours. Quantification of percent age of DRAQ7 cells at 4 hours, quantifed from n=3 –9 positions from 2 separate experiments. **i,** Coating of FITC-CLCs with MPO produced by human peripheral neutrophils following co –incubation with 100µg/ml CLCs for 4 hours. Scale bar **j,** Interaction of dsDNA with Galectin-10 as demonstrated by Biolayer Interferometry (BLI) measurement **k,** Left panel shows an example experiment quantif ying the increase in elastic modulus when human neutrophils are incubat ed with 100µg/ml CLCs in the presence of a hyaluronic acid hydrogel on a cone –and-plate rheometer. Right panel shows the quantification of elastic modulus (G’) at 1Hz at 1 hour (n=2-5 from 4 separate experiments). **l,** An increase in the elastic (G’) modulus of a hyaluoronic acid based hydrogel following incubation with BAL supernatant retrieved from mice 24 hours post-intratracheal administration of PBS, 100µg/mouse CLCs, or 100µg/mouse CLCs + DNAse (n=2-4 mice from 2 experiments) **m,** Immuno-staining of neutrophils with myeloperoxidase (MPO) and Galectin-10 crystals in human mucus plug demonstrating the close interaction of neutrophils with CLCs. Scale bar = 2µm. **n,** Scanning electron microscopy of human neutrophils stimulated with Galectin-10 crystals for 4 hours, demonstrating the penetrance of CLCs by neutrophils and DNA. **o,** Scanning electron microscopy of CLCs incubated with herring sperm DNA (hsDNA) for 4 hours, demonstrating porous holes in crystals **p,** Scanning electron microscopy and **q**, t ransmission electron microscopy of CLCs in a human mucus plug where porous holes in CLCs can be seen.

CLCs have previously been shown to promote neutrophil recruitment and activation^12,21^. Human neutrophils formed NETs in response to CLCs *in vitro*, as evidenced by positive Sytox Green staining (Fig. 3E) and immunolabeling for citrullinated histone H3 (CitH3) and myeloperoxidase (MPO) (Fig. 3F). Both immunostaining and scanning electron microscopy (SEM) revealed extracellular DNA filaments that interconnected multiple crystals, forming web-like structures (Fig. 3F,G). Mouse neutrophils exhibited a comparable response (Supplementary Fig. 3B-E). Real-time live-cell imaging with DRAQ7 staining quantified extracellular DNA release (Fig. 3H; Supplementary Fig. 3F), confirming NET formation over the course of 4 hours. CLCs also induced reactive oxygen species (ROS) production in neutrophils (Supplementary Fig. 3G), consistent with canonical NETosis. Following co-incubation with neutrophils, CLCs became coated with neutrophil-derived proteins (MPO) and extracellular DNA (Fig. 3I, Supplementary Fig. 3H). The apparent interaction of Galectin-10 with DNA was investigated by biolayer interferometry. Galectin-10 interacts with micromolar affinity (*K*_D_ =85 μΜ) with both single– and double-stranded DNA oligonucleotides and via clear association and dissociation binding kinetics (Fig. 3J). This interaction was seen to be independent of the carbohydrate recognition domain (CRD) of Galectin-10 (Supplementary Fig. 3I, and sensitive to elevated salt concentrations, indicating an electrostatically driven interaction (Supplementary Fig. 3J). Given that extracellular DNA substantially contributes to the viscoelastic and tenacious properties of mucus in CF^22^, we next examined how CLC-induced NET formation influences biophysical properties of mucus using a thiol-modified hyaluronic acid hydrogel system^1^. The addition of crystals or neutrophils alone had a minimal impact on either the elastic or viscous modulus of the hydrogel at 1 Hz (Supplementary Fig. 3K-L). In contrast, incubation of CLCs with either mouse or human neutrophils markedly increased the hydrogel’s elastic modulus (Fig. 3K; Supplementary Fig. 3K-M), an effect that was completely abolished by DNase treatment. Similarly, the addition of purified DNA alone recapitulated the increase in elastic, but not viscous, modulus (Supplementary Fig. 3N). Instillation of wild type mice with exogenous CLCs induced robust airway neutrophil recruitment and NET production, peaking at 24 hours post-instillation (Supplementary Fig. 3O). BAL fluid from CLC-exposed mice increased elastic modulus of a hydrogel, whereas this was not seen with BAL fluid from PBS-treated mice or mice co-administered DNase with CLCs (Fig. 3L). These findings demonstrate that DNA released following CLC-induced NET formation both *in vitro* and *in vivo* can significantly affect the biophysical properties of a mucus plug, resulting in more tenacious and pathogenic mucus which contributes to persistence in the airway.

In human samples, neutrophils were frequently observed in close association with CLCs (Fig. 3M). Neutrophil engagement with CLCs *in vitro* induced apparent crystal degradation, seen as superficial holes (Fig. 3N), an effect that was similarly reproduced by extracellular DNA alone (Fig. 3O). Scanning and transmission electron microscopy of patient samples confirmed these porous holes within crystals (Fig. 3P-Q, Supplementary Fig. 3P).

In addition to the release of NETs, which appeared to partially dissolve crystals, neutrophils were observed engulfing CLCs in patient mucus plugs by transmission electron microscopy (Supplementary Fig. 3Q). In mice, neutrophils were confirmed to internalize fluorescently labeled CLCs and accounted for the majority of initial crystal uptake (Supplementary Fig. 3R).

### CLC-containing plugs drive persistent inflammation in vivo

To model CLC-driven airway obstruction in a mouse model, which otherwise lacks the gene for Galectin-10 (*LSGAL10*), CLCs were incorporated into a hydrogel^23^ to mimic mucus and delivered intratracheally to wild type mice.

This model was initially used to develop a translationally relevant study tool which would allow non-invasive detection of CLCs *in vivo* using PET/CT imaging. To this end, a radiolabelled Galectin-10-binding single variable domain on a heavy chain (VHH) antibody lacking CLC-dissolution activity, but with retained CLC binding was generated (Supplementary Fig. 4A-C). Using this radiolabelled VHH, live PET/CT imaging of mice bearing CLC-containing plugs revealed focal lung retention of tracer with a punctate distribution pattern corresponding to patchy plug localization (Fig. 4A). High kidney uptake was consistent with the rapid pharmacokinetic profile of VHHs (Fig. 4B) but a stable and significant retention in the lung indicated a specific binding (Fig 4A-C). *Ex vivo* autoradiography and fluorescence microscopy confirmed tracer colocalization with FITC-labelled CLCs (Fig. 4D). This imaging approach provides a specific, translational tool to visualize mucus plugs laiden with CLCs *in vivo* across muco-obstructive airway diseases.

**Figure 4.**
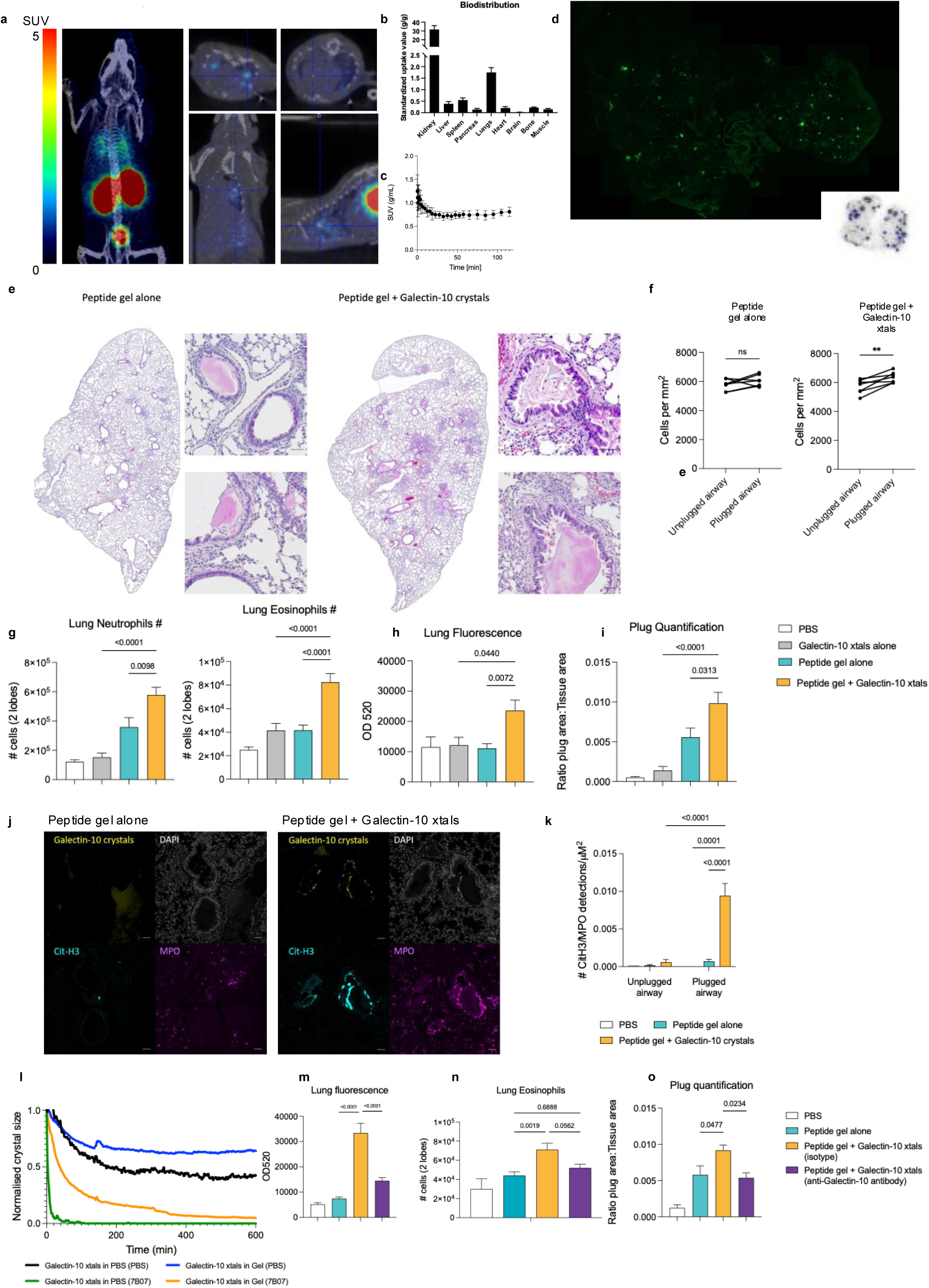
Mucus plugs containing Charcot–Leyden crystals can be reproducible modelled in mice to serve as a therapeutic tool. C57BL/6 mice were intra-tracheally administered PBS, Galectin-10 crystals, Peptide gel alone, or Peptide gel + Galectin-10 crystals. **a,** Example of *in vivo* PET-CT imaging in a mouse instilled with Peptide gel + Galectin-10 crystals and systemically injected with a [89Zr] labelled-VHH to Galectin-10. **b,** Biodistribution of [89Zr] labelled-VHH to Galectin-10 after 2 hours of injection quantified as standardized uptake value (SUV) in mice instilled with Peptide gel + Galectin-10 crystals **c,** Time activit y curves of lungs (Standardized Uptake Values (SUV, g/mL)) 0-120 minutes of [89Zr]Zr-DFO-P01900033 10 in the lungs of mice instilled with Peptide gel + Galectin-10 crystals (n=2). Scan was started 3-7 min p.i. **d,** *Ex vivo* autoradiography of lung slices from mice injected with [89Zr]Zr-DFO-P01900033 demonstrating a similar punctate staining to that acquired by fluorescent (FITC) scanning to detect FITC-labelled Galectin-10 xtals. **e,** Examples of H&E stained lungs from mice administered Peptide gel alone, or Peptide gel + Galectin-10 crystals. Dense cellular infiltration around plugged airways can be clearly seen in the lungs from mice administered Peptide gel + Galectin-10 crystals. **f,** Quantification of number of cells in the 100um radius surrounding plugged or unplugged airways in a cohort of mice administered Peptide gel + Galectin-10 crystals **g,** Flow cytometric quantification of neutrophil and eosinophil inflammation in the lung tissue at 4 days post-administration. **h,** Fluorometric quantificat ion of FITC signal (as a proxy for FITC-labelled Galectin-10 crystals) in the lung tissue at 4 days post-administration. **i,** Quantification of plugs as a ratio of total plug area to total tissue area in H&E stained sections from the lungs of mice at 4 days post-administration. **j,** Immuno-staining of Citrullinated H3 (Cit-H3), Myeloperoxidase (MPO), DAPI and detection of Galectin-10 crystals in mice administered Peptide gel alone or Peptide gel + Galectin-10 crystals 4 days post-administration. **k,** Quantification of the number of CitH3 or MPO or CitH3/ MPO detections as determined by Immuno –staining (f) within the airways of mice administered PBS, Galectin-10 crystals, Peptide gel alone, or Peptide gel + Galectin-10 crystals. **l,** Quantification of crystal size as determined by live imaging over the course of 13 hours incubated in either PBS (Black line) or Peptide gel (Blue line). Additionally, samples are incubated with a Galectin-10 dissolving antibody in either medium, PBS (Green line) or Gel (Orange line) to demonstrate the effi ciency of crystal dissolut ion with antibody, even when embedded in gel. **m-o,** C57BL/6 mice were intra-tracheally administered PBS, Peptide gel alone, or Peptide gel + Galectin-10 crystals, and systemically administered a Galectin-10 dissolving antibody (7B07). **m,** Fluorometric quantification of FITC signal (as a proxy for FITC-labelled Galectin-10 crystals) in the lung tissue at 4 days post-administration. **n,** Flow cytometric quantification of and eosinophil inflammation in the lung tissue at 7 days post-administration. **o,** Quantification of plugs as a ratio of total plug area to total tissue area in H&E stained sections from the lungs of mice at 4 days post-administration.

Next, we studied the biological effects of CLCs within plugs. Administration of gel alone caused transient plugging that resolved within four days, whereas CLC-containing gels produced prolonged obstruction and peribronchial inflammation (Fig. 4E-F), a phenotype strongly resembling the airway plugging seen in humans (Fig 2A-D, 3A-D). The localised inflammation quantified around CLC-plugged airways (Fig 4F) was specified to be both neutrophilic and eosinophilic infiltration (Fig. 4G), with an increased proportion of DCs (Supplementary Fig. 4D). Quantification of fluorescent signal in lung digests verified crystal retention (Fig. 4H), and CLC-containing plugs persisted longer than controls (Fig. 4I). Extensive NET formation was observed within CLC plugs but was absent in unplugged airways, or in CLC-free plugs (Fig. 4J-K). This model faithfully reproduced the human phenotype of mixed eosinophilic-neutrophilic inflammation and demonstated sustained plugging as a result of CLCs.

To test the therapeutic potential of crystal dissolution, mice with CLC-containing plugs were systemically treated with a CLC-dissolving anti-Galectin-10 antibody. The antibody efficiently dissolved CLCs *in vitro*, albeit more slowly within hydrogels than in solution (Fig. 4L). *In vivo*, antibody administration markedly reduced lung crystal burden, inflammatory cell infiltration, and plug persistence (Fig. 4M-O). These findings establish that dissolution of CLCs reverses the key clinical features of CLC-driven airway obstruction.

Thus, in both human disease with ABPA as a model and pre-clinical mouse models, CLCs emerge as key components of mucus plugs. Mechanistially, they are responsible for the recruitment of eosinophils and neutrophils and induction of NETs, which collectively sustain localized inflammatory niches and drive pathogenic mucus plugs. Together, these data identify CLCs as mechanistic drivers and therapeutic targets in muco-obstructive lung disease.

## Discussion

Airway obstruction resulting from mucus plugging is a characteristic yet largely unexplored feature of multiple chronic respiratory diseases, including asthma, COPD, bronchiectasis, CRSwNP, and ABPA^1,3,4,13^. Although mucus plugs have long been viewed as inert byproducts of inflammation, our findings establish them as dynamic inflammatory foci that drive local immune activation, the rendering of pathogenic mucus, and ultimately disease persistence. Effective plug-targeted therapies remain lacking^1,24,25^, emphasising the need to redefine the biology of airway obstruction.

Here, we establish Charcot-Leyden crystals (CLCs) as a defining feature of mucus plugs and subsequent disease. In both human samples and a mouse model we demonstrate that, in contrast to adjacent unobstructed airways, CLC-containing plugs are sites of intense granulocytic inflammation, comprised of both eosinophils and neutrophils. Localised mixed granulocytic inflammation has previously been reported surrounding plugs in fatal asthmatic patients^13,26^, in “non-ventilated” lung segments^27,28^, and in obstructed airways following exacerbations^29^. In asthma, a “mixed granulocytic” phenotype in sputum was associated with low lung function^30,31^, and a severe ventilation defect, and therefore airway obstruction, in the lung^29,32^. Here, we use a pioneering method of imaging-guided bronchoscopy to directly and specifically interrogate plugged airways, uniquely revealing the airway micro-environment localized to obstructed airways.

This study redefines the inflammatory landscape of mucus obstruction. Typically, muco-obstructive diseases in the airways are segregated into neutrophilic (CF, bronchiectasis, COPD) or eosinophilic (asthma) inflammation; however, CLC-containing plugs display both arms of granulocytic inflammation. Even within a patient that could be characterised as an “eosinophilic” endotype based on systemic parameters, neutrophils are seen to infiltrate plugs and produce NETs. This extracelullar extrusion of DNA appears to degrade crystals while simultaneously inducing a more tenacious mucus substance. Based on a sensitivity to salt concentration, the DNA-galectin-10 interaction is likely ionic. Although dimeric Galectin-10 contains two equivalent carbohydrate binding sites which have been shown to bind monosaccharide carbohydrate moieties^12,33^, the broader spectrum of possible molecular partners to Galectin-10 has remained enigmatic^34^. Interestingly, we have observed DNA binding to Galectin-10 carrying a point mutation in its carbohydrate binding site that abbrogates carbohydrate binding. Both uric acid and apatite crystals interact with DNA through electrostatic interactions^35,36^, which have been shown to directly interfere with crystallisation. Furthermore, the addition of highly positively charged peptides to CLCs can result in crystal dissolution *in vitro*^37^. Despite the pronounced eosinophil influx and the established requirement for eosinophil extracellular trap (EET) formation in the generation of CLCs^38^ and in pathogenic mucus in ABPA^39^, these structures were not readily detectable in either our human samples or our mouse models.

Equally, neutrophils were observed to directly engulf CLCs, in both human samples and mouse models. The potential consumption of eosinophils or eosinophil products by neutrophils has been noted surrounding the mucus plugs in patients with fatal asthma^26^. The result of potent neutrophil recruitment or activation in response to CLCs may render these airway plugs as key priming factors for subsequent exacerbations. During a viral exacerbation, the combination of excessive neutrophil influx with pre-existing CLCs could cause expansion of the mucus plugs and even respiratory failure^40^.

Importantly, we demonstrate that the airway niche is fundamentally distinct from systemic compartments. The presence of CLCs, neutrophils, NETs, and even the striking Type-2 inflammatory signature (IL-4, IL-13, GM-CSF) were only detectable in airway samples and not in patient serum (data not shown). Similarly, the accumulation of eosinophil granule proteins in sputum of asthmatic patients in the absence of systemic eosinophilia has been described^41,42^. These data emphasise that representative airway sampling is essential to understand and monitor mucosal disease.

We further establish CLCs a sensitive and specific biomarker of acute ABPA. Airway Galectin-10 levels strongly correlate with CLC burden and are discriminative for acute ABPA, whereas serum Galectin-10 is less performant in capturing this localized pathology. Sputum and BAL fluid provide clinically practical surrogates for plug analysis, but direct retrieval of mucus plugs remains the most informative. The ability of CLC and Galectin-10 as a biomarkers will need to be verified in a broader test population, however, the establishment of enhanced diagnostic criteria for ABPA is vital to enable earlier diagnosis and intervention, preventing airway degeneration, bronchiectasis, and corticosteroid-related complications.

To translate these insights, we developed a Galectin-10-specific PET VHH enabling noninvasive visualization of CLC-containing plugs in vivo. This approach offers a tool for patient stratification and for monitoring responses to therapies aimed at mucus-associated inflammation. CLCs have been sporadically reported in muco-obstructive diseases such as asthma, COPD and CRSwNP^12,43^, but their prevalence and impact on disease severity is still unknown. Using the insights gained from this study, it could be speculated that CLCs are responsible for plug persistence and residual inflammation even with the most advanced treatments. It is increasingly evident that currently available biologics lack penetrance to enable plug resolution^24,25^. The presence of CLCs could render a plug resistant to therapy and a complementary Galectin-10 therapy could allow for remission in these patients. Anecdotally, in this study the persistence of CLCs was observed in patients treated for at least 1 year with omalizumab (n = 3), dupilumab (n = 1) and tezepelumab (n = 1, Fig. 1A-E).

Importantly, we demonstrate that antibody-mediated dissolution of CLCs reduces local inflammation and airway obstruction in a mouse model, demonstrating the therapeutic validity of targeting Galectin-10 in patients with muco-obstructive disease, an approach we are now actively pursuing.

Taken together, these findings position CLCs as central mediators in the pathology of mucus plugs and subsequent disease. CLCs define the persistence, quality and immunologic tone of mucus plugs, while their targeted dissolution represents a rational therapeutic strategy. Future studies should explore the prevalence of CLC-containing plugs across asthma, COPD, bronchiectasis, and CRSwNP, where their presence may underlie poor responsiveness to current biologics. By integrating mechanistic, imaging, and therapeutic approaches, this work reframes mucus plugs as active participants and actionable targets in chronic airway disease.

## Data Availability

All data produced in the present work are contained in the manuscript

## Acknowledgements

We thank the VIB single-cell core, VIB flow core, and VIB-Discovery Sciences for their expert advice and service in this project. We thank MIRaCLe team Leuven for their service and contribution in the establishment of the PET/CT. We thank the lab technicians of the Respiratory Infection and Defense lab, Ann Neesen, Indra De Borle, Anouck Goethals, Katleen De Saedeleer, Greet Barbier and the lab technicians of Bart Lambrecht: Caroline De Wolf, Manon Vanheerswynghels, Kim Deswarte, Karl Vergote and Wendy Touissant for their invaluable contribution to the project. We thank Lotte Terryn for her contributions to the human cytokine work. We thank Sven Verschraegen, Marlies De Coninck (Department of Respiratory Medicine) for their facilitation of pulmonary function testing and the collection of clinical data and patient samples. We thank the referring physicians, CF team and patients. We thank Daan Caudri and Punit Makani (Erasmus MC Rotterdam) for their rendering of the CT scans.

## Materials and methods

### Study Design

The study subjects were recruited in the ABPA Crystal Clear study (BC-09417), a monocentric, prospective cohort study designed to compare patients with and without acute allergic bronchopulmonary aspergillosis (ABPA) in order to identify new biomarkers for ABPA and possible new therapeutic targets. The study was approved by the local ethics committee of Ghent University Hospital (approval number BC-09417), and all participants provided written informed consent in accordance with the 1975 Helsinki Declaration. Samples and data were collected between March 2021 and August 2025 at the Cystic Fibrosis (CF) Reference Centre of Ghent University Hospital, Belgium.

### Patient Cohorts

#### Acute ABPA Patient Cohort

Acute ABPA cases were identified according to the revised ISHAM-ABPA working group criteria with minor adaptations^18^. The case definition of acute ABPA depends on the presence of a history of ABPA. In patients that do not have a history of ABPA, the case definition required: (1) presence of clinical symptoms suggestive of acute ABPA (expectoration of mucus plugs, increased breathlessness, or hemoptysis); (2) sensitization to *Aspergillus* with *Aspergillus*-specific IgE ≥0.35 kU/L; (3) total IgE ≥500 kU/L; and (4) at least 2 of the following 3 criteria: blood eosinophilia ≥500 cells/µL or a history of this, *Aspergillus*-specific IgG ≥72 mg/L (local laboratory reference value, adapted from the original threshold of ≥40 mg/L), or multidetector computed tomography (MDCT) scan abnormalities consistent with acute ABPA (mucus plugging, atelectasis, central bronchiectasis, or high-attenuation mucus). High-attenuation mucus alone was not sufficient to fulfill the case definition, representing a modification from the original criteria. In patients with a history of ABPA, the case definition required: (1) presence of clinical symptoms suggestive of acute ABPA (expectoration of mucus plugs, increased breathlessness, or hemoptysis) and (2) a rise in total IgE >50% from baseline.

Two patients with clinical acute ABPA did not meet this complete case definition and were excluded from ‘acute ABPA’ versus ‘no acute ABPA’ comparisons but included in ‘Charcot-Leyden crystal (CLC) absent’ versus ‘CLC present’ analyses.

The exclusion critera comprise of age <6 years, established diagnosis of eosinophilic granulomatosis with polyangiitis (EGPA), or current treatment with immunosuppressant drugs other than oral corticosteroids.

#### Control Cohorts

Control CF and asthmatic patients were enrolled in parallel to the acute ABPA cohort, including both *Aspergillus fumigatus*-sensitized and unsensitized individuals. No matching was performed; instead, baseline characteristics were compared between groups using appropriate statistical methods.

The exclusion critera comprise of age <2 years, established diagnosis of eosinophilic granulomatosis with polyangiitis (EGPA), current treatment with immunosuppressant drugs other than oral corticosteroids, current non-tuberculous mycobacterial pulmonary disease and unknown sensitization status to *Aspergillus* spp.

#### Cystic Fibrosis Control Cohort

Cystic fibrosis cases were defined as individuals meeting established diagnostic criteria including a positive sweat chloride test (sweat chloride concentration ≥60 mmol/L) and the presence of two CF-causing genetic mutations confirmed by CFTR genotyping. All patients had confirmed CF diagnosis according to international consensus guidelines^44^.

#### Asthma Control Cohort

Asthma cases were required to have a physician diagnosis of asthma based on characteristic symptoms (cough, breathlessness, or dyspnea) combined with demonstration of airflow variability. Airflow variability was confirmed by one or more of the following: an increase in forced expiratory volume in 1 second (FEV1) of ≥12% or 200 mL following inhalation of 400 μg salbutamol, or documented response to inhaled corticosteroids. Alternatively, evidence of airway hyperresponsiveness through methacholine challenge testing was accepted.

The asthma cohort comprised three patient groups: (1) patients who provided informed consent to participate in the Belgian Severe Asthma Registry, (2) patients who were eligible to initiate or switch biological therapies for severe asthma, and (3) asthma patients in whom acute allergic bronchopulmonary aspergillosis (ABPA) was suspected but subsequently excluded.

### Study Procedures and Sample Collection

All patients underwent standardized study procedures: (1) completion of questionnaires as described in section ‘Patient-Reported Outcome Measures’; (2) blood withdrawal for routine laboratory tests and serum storage; (3) collection of spontaneous sputum samples; (4) complete pulmonary function testing (PFT) including FeNO measurement; (5) in a subset of patients, high-resolution multidetector computed tomography (MDCT) scan; in a subset of patients, (6) bronchoscopy (either voluntary or clinically indicated) with collection of mucus plugs, broncho-alveolar lavage fluid (BALF), and bronchial mucosal tissue guided by MDCT results. Underage study participants were not asked to undergo a bronchoscopy for study purposes only. Patients that underwent a bronchoscopy were asked to indicate treatment with inhaled dornase alpha (and if applicable inhaled N-acetylcysteine) on hold for 24 hours and replace it by (hypertonic) saline nebulization. Demographic data (age, sex) and clinical data (active treatment, medical history) were extracted from patient medical records.

For acute ABPA patients, venous blood and spontaneous sputum samples were collected preferably before initiation of specific therapy (oral corticosteroids, itraconazole, or biologicals), representing a state of maximal inflammation. When bronchoscopy was performed (either voluntary or medically indicated), therapy was postponed until after the procedure, and blood samples were collected at the time of bronchoscopy.

#### Pulmonary Function Testing

Spirometry, lung volume measurement, and maximum bronchodilation procedures were conducted according to American Thoracic Society/European Respiratory Society (ATS/ERS) guidelines^45,46^. Total lung capacity (TLC) and residual volume (RV) were measured by body plethysmography.

FeNO was measured using the Niox Vero® (NIOX Inc., North Carolina, USA) electrochemical hand-held analyzer according to ATS/ERS recommendations^47^. FeNO testing was performed before spirometry.

### Patient-Reported Outcome Measures

#### Cystic Fibrosis Questionnaire-Revised (CFQ-R)

Disease-specific health-related quality of life was assessed in CF patients using age-appropriate versions of the Cystic Fibrosis Questionnaire-Revised (CFQ-R), a validated patient-reported outcome measure. For children aged 6-13 years, the CFQ-R Child version was administered as a self-report measure completed by the child, as well as by the parent in a separate questionnaire. For adolescents and adults aged 14 years and older, the CFQ-R Teen/Adult version was used. For this study, we focused on the Respiratory Symptoms domain score as the primary quality of life outcome measure^48^.

#### Quality of Life-Bronchiectasis (QoL-B)

For non-CF patients with bronchiectasis, health-related quality of life was evaluated using the Quality of Life-Bronchiectasis (QoL-B) questionnaire, version 3.1. For this study, we reported the Respiratory Symptoms scale as the primary quality of life outcome measure^49^.

#### Asthma Control Test (ACT)

The ACT is a validated self-administered tool for identifying poorly controlled asthma^50^. The ACT assesses frequency of shortness of breath and general asthma symptoms, use of rescue medications, effect of asthma on daily functioning, and overall self-assessment of asthma control over the previous 4 weeks, rated on a 5-point scale. Scores range from 5 (poor control) to 25 (complete control); a score <20 indicates poor control.

### Bronchoscopy Procedures

#### Patient Selection and Procedure

Bronchoscopy was performed in adult patients and children with clinical symptoms suggestive of ABPA. Controls were selected from patients requiring bronchoscopy with BAL for diagnostic work-up or volunteers with CF. In adults, consent for voluntary bronchoscopy was possible.

Bronchoscopy was performed as a standard outpatient procedure using either a regular flexible bronchoscope (Olympus BF-1TH190; Olympus, Hamburg, Germany) or a thin flexible bronchoscope (Olympus BF-P190; Olympus, Hamburg, Germany). Depending on underlying conditions (excessive bronchial hyperreactivity in young CF patients and ABPA patients), procedures were performed under either procedural sedation and analgesia (PSA) or general anesthesia.

PSA was performed according to established guidelines^51^.Target Ramsay sedation score of 3 was achieved by intravenous titration of midazolam (1 mg/mL, doses 2-10 mg) and fentanyl (0.05 mg/mL, doses 25-50 µg). Supplemental oxygen was administered via nasal canula. When general anesthesia was deemed more appropriate, a standard protocol using intravenous propofol with orotracheal intubation was employed. Topical anesthesia with xylocaine (1 mg/mL) was administered through the bronchoscope in all procedures.

#### Mucus Plug Removal and Sample Collection

The bronchoscope was advanced toward target bronchi containing mucus plugs. Plugs were removed using bronchoscopic suction; in some cases, forceps were used to remove small pieces. After plug removal, a 20 mL bronchial lavage was performed to remove residual viscous material. Mucus plugs obtained via bronchoscopy were used for processing into supernatant and cell fractions with consequent cytological diagnosis and formalin fixation and paraffin embedding.

#### Bronchial Mucosal Biopsies

Mucosal biopsies were obtained using both conventional forceps (single use biopsy forceps, ø2.3mm, Micro-Tech (Nanjing), China) and cryotechnique (flexible cryoprobe, ø 1.7 mm; Erbe, Tübingen, Germany)^52^. Mucosal cryobiopsies were obtained from longitudinal stretches at subsegmental levels at the operator’s discretion. The cryoprobe was placed adjacent to the airway wall before initiating a 2-3 second freeze cycle, after which the probe and bronchoscope were gently retracted together. Biopsies were retrieved from both plugged bronchi and contralateral anatomically equivalent unplugged bronchi. After removal, biopsies were either stored in liquid nitrogen for later fixation (overnight in 4% buffered formaldehyde) and paraffin embedding, or embedded directly in Optimal Cutting Temperature (OCT) compound for cryosectioning.

### Sample Processing and Storage

All collected samples were processed on the day of collection and stored either in the ‘Prospective Biobank CF, Bronchiectasis, Respiratory Infections’ (BR151, BC-08261, B.U.N. 6702020001009, FAGG reference number BB210012) or the ‘Prospective Biobank for asthma and COPD’ (BR-21, BC-05210, B.U.N. 670201940238, FAGG reference number BB190132). Unique patient samples were aliquoted to prevent degradation from repeated freeze-thaw cycles and maintain specimen integrity.

#### Serum

Fresh blood was collected in serum tubes and centrifuged at 20°C for 10 minutes at 400 g. Cell-free serum was aliquoted and stored at –80°C until analysis.

#### Sputum and Mucus Plugs

Spontaneous sputum and bronchoscopically removed mucus plugs were collected in sterile recipients and processed within 3 hours after collection as follows. After separating saliva from mucus and weighing the sample, specimens were incubated with dithiothreitol (DTT) solution, vortexed, and rocked for 15 minutes at 37°C. An equal volume of phosphate-buffered saline (PBS) was added, samples were vortexed for 15 seconds and rocked for 5 minutes. Samples were then filtered and centrifuged. Cell-free supernatant was aspirated and stored at –80°C. The cell suspension was used to prepare cytospins at 50,000 cells per slide.

#### Bronchoalveolar Lavage Fluid

During bronchoscopy, 2 aliquots of 20 mL prewarmed sterile 0.9% sodium chloride solution were instilled and re-aspirated after collection of samples for clinical purposes. When possible, samples were collected from formerly plugged lung segments (after plug removal). The aspirated BAL fluid was immediately placed on ice and processed. Fluid was filtered and centrifuged, and cell-free supernatant aliquots were stored at –80°C. The cell suspension was used to prepare cytospins at 50,000 cells per slide.

#### Cell Counting and Cytospin Preparation

Airway samples (BALF, spontaneous sputum, and bronchoscopically removed mucus) were processed to obtain cytospins for cell and CLC counting. PBS was added to cell pellets remaining after supernatant aspiration. For cell counting, 10 µL cell suspension was mixed with 40 µL Türk’s solution, and 25 squares were counted on a Bürker hemocytometer. For differential cell counts, slides were prepared with 50,000 cells per slide on Superfrost Plus slides and stained with Epredia Shandon Kwik-Diff (9990700) stain. For CLC counting, cytospins were scanned using an Axio Scan Z1 device (Carl Zeiss, Germany), and CLCs were manually annotated using QuPath software.

### Biomarker Assays

#### Galectin-10 Detection

Galectin-10 concentrations were measured using an in-house sandwich enzyme-linked immunosorbent assay (ELISA). Nunc Maxisorp 96-well plates (catalog 469949, Thermo Fisher Scientific) were coated with capture antibody \ in PBS and incubated overnight at 4°C. Plates were washed five times with PBS containing 0.05% Tween-20 (PBST) and blocked with proprietary blocking buffer for 2 hours at room temperature.

Following five washes with PBST, serum samples and sputum/plug supernatants were diluted in proprietary diluent were added, 50 μL/well, and incubated for 2 hours at room temperature. Recombinant His-Tagged galectin-10 standards (VIB protein core, Ghent, Belgium) were prepared in diluent to generate a calibration curve ranging from 0.1 to 25 ng/mL in eight standard points with 2.5 fold serial dilutoions. After five washes, biotinylated detection antibody was added and incubated for 1 hour at room temperature. Plates were washed five times with PBST, followed by addition of streptavidin-horseradish peroxidase (SA-HRP) (Jackson ImmunoResearch, 016-030-084) at for 30 minutes at room temperature. After five final washes, 3,3’,5,5’-tetramethylbenzidine (TMB) (Sigma-Aldrich, CL07-1000mL) substrate, 50 μL/well, was added and the reaction was stopped with 0.5M H_2_SO_4_, 50 μL/well. Absorbance was measured at 450 nm with 650 nm reference wavelength subtraction using a Victor Nivo microplate reader (PerkinElmer). The dynamic range was set at 0.3-4 ng/mL, with a lower limit of quantification (LLOQ) of 0.3 ng/mL and a higher limit of qauantification (HLOQ) of 4 ng/mL. When measurements were below the LLOQ, a value was given of LLOQ*dilution/2. When measurements were above HLOQ, a value was given of HLOQ*dilution. Samples were stored in Eppendorf tubes at –80°C prior to analysis. Samples were run in duplicate, or triplicate where technically feasible. Specific antibody clones and concentrations remain proprietary.

#### Cytokine and Chemokine Quantification

Serum cytokines (IFN-γ, IL-1β, IL-2, IL-4, IL-5, IL-6, IL-8, IL-10, IL-13, IL-17A, TNF-α, GM-CSF) were quantified using Meso Scale Discovery V-PLEX Proinflammatory Panel 1 Human Kit and V-PLEX Cytokine Panel 1 Human Kit (Meso Scale Discovery K15049D-1 and K15050D-1) according to the manufacturer’s protocol, with data acquired on a MESO QuickPlex SQ 120. For values below the lower limit of quantification (LLOQ), LLOQ/2 was used for statistical analysis. For values exceeding the upper limit of quantification, the upper limit value was used.

Spontaneous sputum and mucus plug supernatant cytokines (IFN-γ, IL-1β, IL-2, IL-4, IL-5, IL-6, IL-8, IL-10, IL-13, IL-17A, TNF-α, GM-CSF) were quantified using the same V-PLEX kits. Samples were diluted 3-fold in PBS containing 0.5% bovine serum albumin. TSLP and IL-9 were quantified using V-PLEX Cytokine Panel 2 Human Kit (K15084D-1) with 2-fold sample dilution [18]. Chemokines (eotaxin-1, eotaxin-3, IP-10) were quantified using V-PLEX Chemokine Panel 1 (human) Gen. B Kit (K0082723) with 2.5-fold sample dilution.

#### Neutrophil Elastase-DNA Complex ELISA

Neutrophil extracellular trap (NET) formation was assessed by quantifying neutrophil elastase (NE)-DNA complexes using an in-house enzyme-linked immunosorbent assay (ELISA)(adapted from^53^). Nunc Maxisorp 96-well plates (catalog 469949, Thermo Fisher Scientific) were coated with 50 µL per well of mouse monoclonal anti-neutrophil elastase antibody (clone SC-55549, Santa Cruz Biotechnology; original clone SC-9521) diluted 1:250 in coating buffer (15 mM Na₂CO₃, 35 mM NaHCO₃, pH 9.6) and incubated overnight at 4°C.

Plates were washed three times with 200 µL per well PBS and blocked with 200 µL per well of 5% bovine serum albumin (BSA) in PBS for 2 hours at room temperature. After three washes with PBS (200 µL/well), samples were added at 50 µL total volume in PBS containing 1% BSA and incubated for 2 hours at room temperature. Sputum and plug supernatant samples were added at 10 µL per well with 40 µL of PBS/1% BSA. A serial 1:5 dilution series was included as standards. Control wells included PBS blanks (n=2), secondary antibody-only controls (n=2), and no-primary antibody controls (n=2).

Following three washes with 300 µL per well wash buffer (PBS/1% BSA/0.05% Tween-20), plates were incubated with 50 µL per well anti-DNA-peroxidase antibody (from Cell Death Detection ELISA kit, catalog 11774425001, Roche) as per manufacturer’s instructions and optical density was measured at 405 nm using a Infinite® M-plex (Tecan).

### Histological and Imaging Methods

#### Histochemical Staining

Spontaneous sputum and mucus plugs were fixed and embedded in paraffin, sectioned at 3 µm, and stained with hematoxylin and eosin (H&E) and Masson’s trichrome according to standard protocols. Bronchial mucosal biopsies (both cryobiopsies in OCT and conventional biopsies in 4% paraformaldehyde with paraffin embedding) were stained with H&E and Congo red for eosinophil identification and quantification.

### Immunofluorescence Staining

#### Sputum and Mucus Plug Immunofluorescence

Paraffin-embedded sputum and mucus plugs were prepared and stained with antibodies against myeloperoxidase (MPO) (Abcam ab225474), galectin-10 (Gal-10) (R&D Systems AF5447), and *Aspergillus* (Abcam ab20419), with DAPI nuclear counterstain. All stainings are isotype controlled.

#### Tissue Preparation and Sectioning

Human tissue specimens were fixed in 4% neutral buffered formalin for 24-48 hours at 4°C on a rotor, dehydrated through graded ethanol series (70%, 80%, 96%, and 100%), cleared in xylene, and embedded in paraffin wax according to standard histological procedures. Serial 3-µm-thick sections were cut using a microtome, floated in a 60°C water bath, and mounted on positively charged glass slides (SuperFrost Plus, Thermo Fisher Scientific). Slides were dried overnight at 37°C and stored at room temperature until use.

#### Deparaffinization and Rehydration

Tissue sections were deparaffinized and rehydrated using the following protocol: xylene for 10 minutes (two changes), 100% ethanol for 5 minutes (two changes), 95% ethanol for 5 minutes, 70% ethanol for 5 minutes, and distilled water for 5 minutes. All steps were performed at room temperature with gentle agitation.

#### Antigen Retrieval

Heat-induced epitope retrieval (HIER) was performed to unmask antigens and reduce autofluorescence. Slides were immersed in citrate buffer (10 mM sodium citrate, pH 6.0), and heated in a pressure cooker for 10 minutes or in a water bath at 90°C for 45 minutes. After antigen retrieval, slides were cooled to room temperature for 20-30 minutes and washed three times in phosphate-buffered saline (PBS, pH 7.4) for 5 minutes each.

#### Blocking and Antibody Incubation

Non-specific binding was blocked by incubation with blocking buffer (0.3% Triton X-100 in PBS) for 30 minutes at room temperature in a humidified chamber. Sections were then incubated with primary antibodies diluted in antibody dilution buffer (1% BSA in PBS) for 1 hour at room temperature. Primary antibody concentrations were optimized for each antigen.

The following primary antibodies were used: anti-myeloperoxidase (MPO) (directly conjugated to Alexa Fluor 488, Abcam ab225474), anti-galectin-10 (Gal-10) (R&D Systems AF5447), and anti-*Aspergillus* (Abcam ab20419), with DAPI nuclear counterstain. Isotype-matched control antibodies were used as negative controls.

After primary antibody incubation, slides were washed three times in PBS containing 0.1% Triton X-100 (PBST) for 5 minutes each. Sections were then incubated with species-appropriate Alexa Fluor-conjugated secondary antibodies (Thermo Fisher Scientific) diluted 1:500-1:1000 in antibody dilution buffer for 1 hour at room temperature in the dark. Following secondary antibody incubation, the slides were incubated with the anti-MPO antibody direrctly conjugated to Alexa Fluor 488. Finally, slides were washed three times in PBST for 5 minutes each in the dark.

#### Nuclear Counterstaining and Mounting

Nuclei were counterstained with 4′,6-diamidino-2-phenylindole (DAPI; 1 µg/mL in PBS) for 5 minutes at room temperature, followed by three washes in PBS for 5 minutes each. Coverslips were mounted using ProLong Diamond antifade mounting medium (Thermo Fisher Scientific). Slides were allowed to cure overnight in the dark at room temperature or for 24 hours at 4°C before imaging.

#### Image Acquisition

Fluorescence images were acquired using an automated slide scanner (Zeiss Axio Scan 7, Carl Zeiss) equipped with appropriate laser lines and filter sets for Alexa Fluor 488, 555, 647, and DAPI. Images were captured using a 20× objective with sequential scanning to minimize spectral bleed-through. For each sample, multiple fields of view were acquired with consistent exposure settings.

#### Bronchial Mucosal Biopsy Immunofluorescence

Paraffin-embedded bronchial mucosal biopsies were stained using the same antibodies (anti-MPO, anti-Gal-10) and protocol as described above for sputum and plug samples, with DAPI nuclear counterstain.

#### Human/Mouse neutrophil NET immunofluorescence

After 4 hours of stimulation in a µ-slide 8 well 3D ibiTreat (iBidi) neutrophils were fixed in 4% PFA (Boster Biological Technology) for 10 minutes at room temperature. Cells were gently washed, and blocked in PBS, 2% Bovine Serum albumin (Sigma Aldrich) and 5% Donkey serum (Sigma-Aldrich) for one hour at RT before being incubated with primary antibodies overnight at 4°C. Primary antibodies consisted of anti-MPO (AF3667-SP R&D), anti-CitH3 (Abcam ab5103). Cells were gently washed and stained for fluorescence with AF647 anti-Goat IgG (A-21447 Thermofisher) and Cy3 anti-Rabbit IgG (711-166-152 Jackon ImmunoResearch) for 1 hour at RT in the dark. Nuclei were counterstained with DAPI (ThermoFisher Scientific) for 15 minutes and imaged on a Zeiss LSM880 Microscope.

### Image Analysis and Quantification

#### Quantification of eosinophils in bronchial biopsies

All histological and immunofluorescence images were analyzed using Zen Blue (version Zen 3.13) and QuPath software (version 0.6.0). For eosinophil quantification in bronchial biopsies, Congo red-stained sections were manually counted. CLC quantification was performed on Kwik-Diff™ stained and scanned cytospin slides with manual annotation.

#### Manual quantification of inflammation and NETs

Using QuPath V0.5.1 airways were manually annotated as plugged or unplugged. Cell detection and segmentation was performed based on DAPI signal. Within these regions, CitH3 or MPO or CitH3/MPO was detected and total area of detections was consolidated per mouse.

### Animal experiments

#### Animals

Wildtype C57BL/6 mice were purchased from Janvier Labs (Saint-Berthevin, France). Mice were housed under specific pathogen-free conditions and used between 7 and 12 weeks of age. All experiments were approved by the animal ethics committee at Ghent University (EC2023-2024, EC2025-078) and were in accordance with Belgian animal protection law.

#### Preparation of Charcot-Leyden crystals

Recombinant crystals were prepared as in Persson et al^12^

#### Preparation of Hyaluronic acid gel

Glycosil Thiolated Hyaluronon (GS222F-5EA Advanced Biomatrix) was dissolved in 2ml of ddH_2_O on a rotator for 2 hours. PEGSSDA (GS755-2EA Advanced Biomatrix) was dissolved in 0.5ml of degassed ddH_2_O and 270µl of PEGSSDA was incubated with 2ml of Glycosil overnight at RT.

#### Preparation of Peptide gel

CM63 peptide (H-FQFQFK-NH2 Ontores) (A kind gift of Richard Hoogenboom) was dissolved in 0.9x PBS to give a 1% w/v gel. This was dissolved with extensive vortexing and left to gel at RT overnight^23^.

#### Administration of gel or CLCs to mice

Peptide hydrogel 1% (w/v) was prepared as described. PBS or FITC-labelled Gal10 crystals were added in a 1/10 dilution to peptide gels to a final concentration of 1.43 Galectin-10 xtals mg/mL. In total 100 µg Gal10 crystals was administered in 70 µl of 1% peptide hydrogel, PBS or PBS containing 100 U/ml DNase I (Roche) via the intratracheal route to 7 week old male C57BL/6 mice, anesthetized lightly with isoflurane (2 liters/min, 2 to 3%; Abbott Laboratories). Mice were monitored daily for weight, appearance and behavior.

#### Processing of mouse lungs for fluorescence, flow cytometry and histology

Mice were euthanized by an overdose of pentobarbital (KELA Laboratoria) and lungs were collected. The middle right, inferior right lobe and the left lobe of the mouse lung were removed from the mouse without inflation and washed in PBS before overnight fixation in 4% PFA (AR1068 – Boster Biological Technology) at 4°C.

The superior and post-caval lobe were isolated and minced finely with scissors and then digested for 30 minutes in RPMI-1640 (Gibco, Thermo Fisher Scientific) containing 20 μg/ml liberase TM (Roche), 10 U/ml DNase I (Roche), and 5% of FCS (Bodinco) at 37°C. 300μl of this 2ml lung digestion mix was removed with a cut pipette tip after extensive vortexing and placed into a black 96-well flat clear bottomed plate (Greiner). Plate was spun at 400g for 3 minutes and fluorescence was acquired with a 480/520 emission/excitation using an Infinite® M-plex (Tecan). Absorbance from 410nm to 710nm was acquired and averaged and used as reference wavelength to adjust fluorescent signal to.

The remaining lung digest was processed into a single cell suspension for flow cytometry. Lungs were filtered through a 70 μm cell strainer and were depleted of red blood cells (RBCs) by using RBC lysis buffer [0.15 M NH4Cl (Merck), 1 mM KHCO3 (Merck), and 0.1 mM Na2-EDTA (Merck) in MilliQ H2O] produced in-house. Single-cell suspensions were incubated with a mix of fluorescently labelled monoclonal antibodies (Ab) in the dark for 30 min at 4°C. To reduce non-specific binding, 2.4G2 Fc receptor Ab (Bioceros) was added. Dead cells were removed from analysis, using fixable viability dye eFluor780 (Thermo Fisher Scientific). 123count eBeads Counting Beads (Thermo Fisher Scientific) were added to each sample to determine absolute cell numbers. Before acquisition, photomultiplier tube voltages were adjusted to minimize fluorescence spillover. Single-stain controls were prepared with UltraComp eBeads (Thermo Fisher Scientific) following the manufacturer’s instructions and were used to calculate a compensation matrix. Sample acquisition was performed on a LSR Fortessa cytometer equipped with FACSDiva software (BD Biosciences). Final analysis and graphical output were performed using FlowJo software (Tree Star, Inc) and GraphPad Prism 10 (GraphPad Software, Inc). The following antibodies were used: anti-CD125 (BV421, BD Biosciences), anti-CD64 (BV711, BioLegend), anti-Siglec F (BV786, BD Biosciences), anti-CD19 (APC, BD Biosciences), anti-CD3 (APC, BioLegend), anti-Ly6C (AF700, BD Biosciences), anti-MHCII (Pe, Thermo Fisher Scientific), anti-CD45 (Pe-Cy5, BioLegend), anti-CD11b (Pe-Cy7, BD Biosciences), anti-Ly-6G (BUV395, BD Biosciences), anti-CD11c (BUV737, BD Biosciences).

#### Histology and quantification

Lungs were embedded in paraffin and 6µm slices were taken at 3 depths with 150µM distance and placed on the same slide. Slides were stained with hemotoxylin and eosin and scanned with an Zeiss AxioScan Z1. To quantify plugs, images were analysed with QuPath V0.5.1. Colour deconvolution was performed, and multiple images were consolidated into a training image, and “Plugs” and “Not plugs” were manually annotated to train a pixel classifier. Tissue detection and plug detection were run on each image and the ratio of plug to total tissue area was plotted, and averaged across the 3 lobes over a total of 3 lung depths (9 lobes in total).

For immunostaining, lungs were dewaxed using an autostainer (Leica) and incubated in a pressure cooker (Retriever 2100) in DAKO solution (Agilent Technologies Belgium). Samples were washed in PBS, blocked in 2% Bovine Serum albumin (Sigma Aldrich) and 5% Donkey serum (Sigma-Aldrich) for one hour at RT before being incubated with primary antibodies overnight at 4°C. Primary antibodies consisted of anti-MPO (AF3667-SP R&D), anti-CitH3 (Abcam ab5103). Cells were gently washed and stained for fluorescence with AF647 anti-Goat IgG (A-21447 Thermofisher) and Cy3 anti-Rabbit IgG (711-166-152 Jackon ImmunoResearch) for 1 hour at RT in the dark. Nuclei were counterstained with DAPI (ThermoFisher Scientific) for 15 minutes, slides were mounted with Polyvinyl alcohol (Sigma) and imaged using an Axioscan 7 (Zeiss).

#### PET/CT Tracer

Gal10-binding VHHs were described previously (WO2019197675). The sequences of Gal10 VHHs were recloned as untagged or C-terminal His₆-tagged variants for production in in E. coli, with framework residues standardized for stability. VHHs were purified from the periplasm by IMAC on Ni Sepharose) or by Protein A for tagless variants, followed by desalting and size exclusion chromatography (SEC). The purity and integrity were confirmed by SDS-PAGE, mass spectrometry (MS), and endoxin levels in a LAL assay. The CLC dissolution capacity of Gal10 VHHs was monitored by spinning disk microscopy (Zeiss Axio Observer.Z1) in comparison to Gal10 1D11 Fab (35-60 µM) and controls (n=3). CLCs generated from recombinant Gal10 were spotted in 96-well plates and imaged every 3-5 min for 3h. CLC area was quantified in ImageJ, normalized to t₀, and EC₅₀/EC₁₀ values derived by nonlinear regression (GraphPad Prism).

For 89Zr immuno-positron emission tomography (PET) imaging studies, VHHs were derivatized to a octadentate bifunctional chelate desferrioxamine (DFO*NCS), essentially according to the protocol of Vugts et al. 2017^54^. Two VHHs (a CLC dissoluting, and a non-CLC dissoluting VHH) were humanized to IGHV3/JH consensus sequence, and removal of post-translational modification sites to improve thermal stability. Sequence optimized and humanized VHHs P01900033 and P01900038 were derivatised to DFO*-NCS (ABX Chemicals) in ten-fold molar excess in 50 mM NaHCO₃ buffer, followed by purification by SEC. The degree of incorporation of DFO* was assessed by MS analysis. In the DFO-P01900033 a fraction of 67% fraction contained one DFO moiety, for DFO-P01900038 a 8% fraction had one DFO group incorporated. The Gal10 binding affinities of the cold-labelled DFO*VHH conjugates and reference compounds was confirmed with BLI.

Radiolabeling with Zirconium-89 (⁸⁹Zr) in 1 M oxalic acid (Cyclotron VU, Amsterdam, Netherlands) was done on 5-10 nmol of DFO*conjugate in 0.5 M HEPES (pH 7.2, 37 °C, 1 h), and purified by Amicon ultrafiltration with a 10 kDa MWCO membrane (Sigma-Aldrich) into PBS/ascorbic acid (5 mg/mL) formulation buffer. The VHH concentration was supplemented with unlabeled VHH to obtain a final concentration of 20 nmol/mL. The final tracer stock solution contained 20.7 MBq/mL activity with a radiochemical purity >99%, assessed by radio-SEC-HPLC prior to in vivo injection

For µPET/CT, tracers (20 MBq, 20 nmol VHH) were injected intravenously ([⁸⁹Zr]Zr-DFO*-P01900033: 2.07 ± 0.09 MBq/1.66 ± 0.07 nmol; [⁸⁹Zr]Zr-DFO*-P01900038: 2.2 ± 0.14 MBq/1.98 ± 0.13 nmol). Dynamic µPET/CT (β-cube/X-cube, Molecubes, Belgium) was acquired for 2 h, followed by ex vivo biodistribution and lung histology. Serum radiometabolites were analyzed 10 min post-injection (n=3). CLC binding was assessed by autoradiography on mouse lung cryosections (n=4/group).

#### Antibody administration

24 hours before administration of Peptide gel, mice were administered 2mg of 7B07 (an anti-Galectin-10 antibody derived from a llama immunization campaign towards the crystalline form of human Galectin-10. It was characterized as fast and potent CLC dissolving lead) or isotype control (Synagis/Palivizumab (AstraZeneca) in 200µl via intravenous injection in the tail vein.

### *Ex vivo* Neutrophil experiments

#### Human neutrophil isolation

Human blood was collected from healthy donors (Ethical approval by UZ Gent under (056-2019/SDS)) in K-EDTA tubes and processed immediately. 7ml of blood was gently layered on 7ml of Polymorph prep (Fisher Scientific) and centrifuged for 35 minutes at 500g at RT without a break. The PMN layer was collected, washed and residual red blood cells were lysed with 1x lysis buffer (eBioscience). Neutrophils were counted and resuspended at 5 x 10^6^ cells/ml and stimulated with 100ng/ml GM-CSF (Peprotech) for 30 minutes before stimulation with crystals. Neutrophils were stimulated with 100µg/ml of FITC-labelled Galectin-10 crystals for 4 hours. Extracellular DNA was stained with 1µM Sytox green (Thermofisher). ROS production was measured using luminol assay. Briefly, neutrophils were resuspended in HBSS at 2 x 10^6^ cells/ml with HEPES (20mM), CaCl2 (1 mM), MgCl2 (0.5 mM), HRP (Sigma, 8 U/ml) and luminol (Sigma, 0.1 mM). Cells were immediately plated (100µl) into a white 96 well plate (Greiner) containing PMA, soluble Galectin-10 or Galectin-10 crystals. Luminescence was measured every minute.

#### Mouse neutrophil isolation

Bone marrow of mice femurs and tibias were flushed cells were separated via density centrifugation using a gradient of Histopaque 1119 and Histopaque 1077 (Sigma Aldrich) (Detailed protocol Wishart et al^55^). Residual red blood cells were lysed with 1x lysis buffer (eBioscience. Neutrophils were counted and resuspended at 5 x 10^6^ cells/ml and stimulated with 100ng/ml GM-CSF (generated at VIB) for 30 minutes before stimulation with crystals (see Persson et al for preparation) for 4 hours. Extracellular DNA was stained with 1µM Sytox green (Thermofisher).

#### In vitro dissolution of crystals

Crystals at 0.7mg/ml were deposited on the bottom of an 8 well 3D ibiTreat (iBidi). 1% peptide gel (or PBS) was gently deposited on top of the crystals and allowed to equilibrate for 30 minutes. 20µM of 7B07 was pipetted on top of the solidified gel and images were captured with a Spinning disk (Zeiss) at 5x magnification. Extended depth of focus were created from the stacks, on which crystal size was measure over time, and normalized to size at time point 0 (ImageJ).

### Rheology

All rheological measurements were performed on a DHR2 (TA Waters) using a heated peltier plate at 37°C, a 40,0mm 0,4925° cone, and a solute trap. 250ul of hyaluronic acid hydrogel was loaded onto the peltier plate at and allowed to equilibrate for 5 minutes. 250,000 neutrophils were centrifuged and resuspended in 25ul PBS or 25ul PBS containing 1mg/ml Galectin-10 crystals. Neutrophils were mixed with hydrogel on the peltier plate and incubated for 60 minutes oscillating at 1 Hz and 5% strain with a solute trap.

### Electron Microscopy

#### Scanning Electron Microscopy – Plugs

Sputum plugs were collected in 2 mL Eppendorf tubes and fixed overnight at 4°C in 2% paraformaldehyde and 2.5% glutaraldehyde in 0.1 M sodium cacodylate buffer. Samples were washed three times in buffer and post-fixed in 1% reduced OsO_4_ in 0.1 M sodium cacodylate buffer for 1 hour on ice. After three washes in distilled water, samples were dehydrated through a graded ethanol series, followed by an ethanol:acetone (1:1) step and final incubation in 100% acetone. Dehydrated specimens were critical point dried (Leica EM CPD300), mounted on aluminum stubs with carbon adhesive tape, and sputter-coated three times with 5 nm platinum (Quorum Q150T ES) to minimize charging. Imaging was performed on a Zeiss Crossbeam 540 scanning electron microscope at 1.5 kV using the secondary electron (SE) detector.

#### Scanning Electron Microscopy – Neutrophils

For scanning electron microscopy (SEM), samples were collected in 1,5ml Eppendorf tubes and fixed at room temperature by adding an equal volume of fixative (4% paraformaldehyde and 2.5% glutaraldehyde in 0.1 M sodium cacodylate buffer). After 10 min, the cells were carefully centrifuged for 2 min at 300 × g. The supernatant was removed and replaced with 500 µl of fixative, followed by gentle rotation for 1 h at room temperature.

After fixation, the cells were centrifuged again and washed three times in 0.1 M sodium cacodylate buffer. They were post-fixed in 1% reduced osmium tetroxide (OsO₄) in 0.1 M sodium cacodylate buffer for 1 h on ice. Following three washes in ultrapure water, the samples were dehydrated through a graded ethanol series (30%, 50%, 70%, 95%, 100% EtOH) with gentle rotation for 15 min per step.

After the final dehydration step, an equal volume of acetone was added. After 15 min and light centrifugation, the solution was replaced by 100% acetone. Subsequently, 5 µl of each sample was blotted onto a silicon wafer mounted on aluminum stubs with carbon adhesive tape. After air drying, the samples were sputter-coated twice with 5 nm platinum (Quorum Q150T ES) to minimize charging.

Imaging was performed using a Zeiss Crossbeam 540 scanning electron microscope operated at 1.5 kV with a secondary electron (SE) detector.

#### Transmission Electron Microscopy

Sputum plugs were immediately immersed in fixative (4% formaldehyde, 2.5% glutaraldehyde in 0.1 M sodium cacodylate buffer) and cut into 1 mm³ fragments, which were transferred to 2 mL Eppendorf tubes. Samples were placed in a vacuum oven for 30 minutes and subsequently rotated for 3 hours at room temperature. The fixative was then replaced with fresh solution, and samples were rotated overnight at 4°C. After rinsing, specimens were post-fixed overnight at 4°C in 1% reduced OsO_4_ in 0.1 M sodium cacodylate buffer, washed three times, and contrasted with 1% uranyl acetate in distilled water for 1 hour at 4°C. Following additional washes in distilled water, samples were dehydrated through a graded ethanol series and embedded in Spurr’s resin. For crystal detection, semithin sections (0.5 µm) were prepared with an ultramicrotome (Leica EM UC7) and stained with toluidine blue. After screening, ultrathin sections (70 nm) were collected on Formvar-coated copper slot grids and post-stained with 1% uranyl acetate (40 minutes) and 3% lead citrate (10 minutes) using a Leica EM AC20 automated stainer. Sections were examined with a JEM-1400Plus transmission electron microscope (JEOL, Tokyo, Japan) operating at 80 kV.

### Live cell imaging

2 x 10^4^ neutrophils were plated in a µ-slide 15 well 3D ibiTreat (iBidi) and incubated with DRAQ7 (ThermoFisher Scientific) (1/5000) and Hoechst (Hoechst 33342, ThermoFisher Scientific) (1/1000). Image analysis was performed using arivis Vision4D (version 4.1.2, arivis AG, Germany) to segment and quantify neutrophil extracellular traps (NETs) based on combined intensity and size thresholding. The Blob Finder tool was applied to the Hoechst channel to identify nuclei, while a simple intensity threshold was applied to the DRAQ7 channel to detect NET structures. NETs were classified as early-stage when segmented objects measured <150 voxels.

### Biolayer Interferometry (BLI)

BLI experiments were performed in PBS-buffer supplemented with 0.1% (w/v) BSA and 0.02% (v/v) Tween 20, with an Octet RED96 instrument (FortéBio), or in indicted increasing salt concentrations of NaCl operating at 25 °C. Streptavidin-coated biosensors were functionalized with biotinylated ssDNA or dsDNA (CTCGACGATAAAAATCGCCCGCGCCGCCCGCGCTCCTCGGATAAATATCTA) and then dipped into solutions containing different analyte concentrations. His-Tag Galectin-10 WT or CRD (W72A mutation) were used as analyte (see Persson et al, 2019 for details of Galectin-10 production). To verify that no non-specific binding was present during the interaction assay, non-functionalized biosensors were used as a control. The sensor traces from zero concentration samples were subtracted from the raw data traces before data analysis. To correct for bulk effects during the measurements for the interaction between DNA and Galectin-10 a column of non-functionalized sensors was used to enable double reference subtraction. All data were fitted with the FortéBio Data Analysis 9.0.0.4 software using a 1:1 ligand model.

### Statistical Analysis

Statistical analyses were performed using R version 4.3.1 (R Foundation for Statistical Computing) and Prism version 10.5.0 (GraphPad Software, San Diego, California). The following R packages were used: readxl (version 1.4.3) for importing data from Excel files, dplyr (version 1.1.4) for data manipulation and transformation, tidyr (version 1.3.1) for data tidying and reshaping, ggplot2 (version 3.5.1) for data visualization and generation of publication-quality figures, and scales (version 1.3.0) for scale functions and formatting. Biomarker values were analyzed using nonparametric tests as implemented in base R: unpaired Mann-Whitney *U* test or paired Wilcoxon matched-pairs signed-rank test for two-group comparisons. Results with *P* < 0.05 were considered significant. Significance levels were denoted as: *, *P* < 0.05; **, *P* < 0.01; ***, *P* < 0.001; and ****, *P* < 0.0001.”

---

**Supplementary Table 1.**
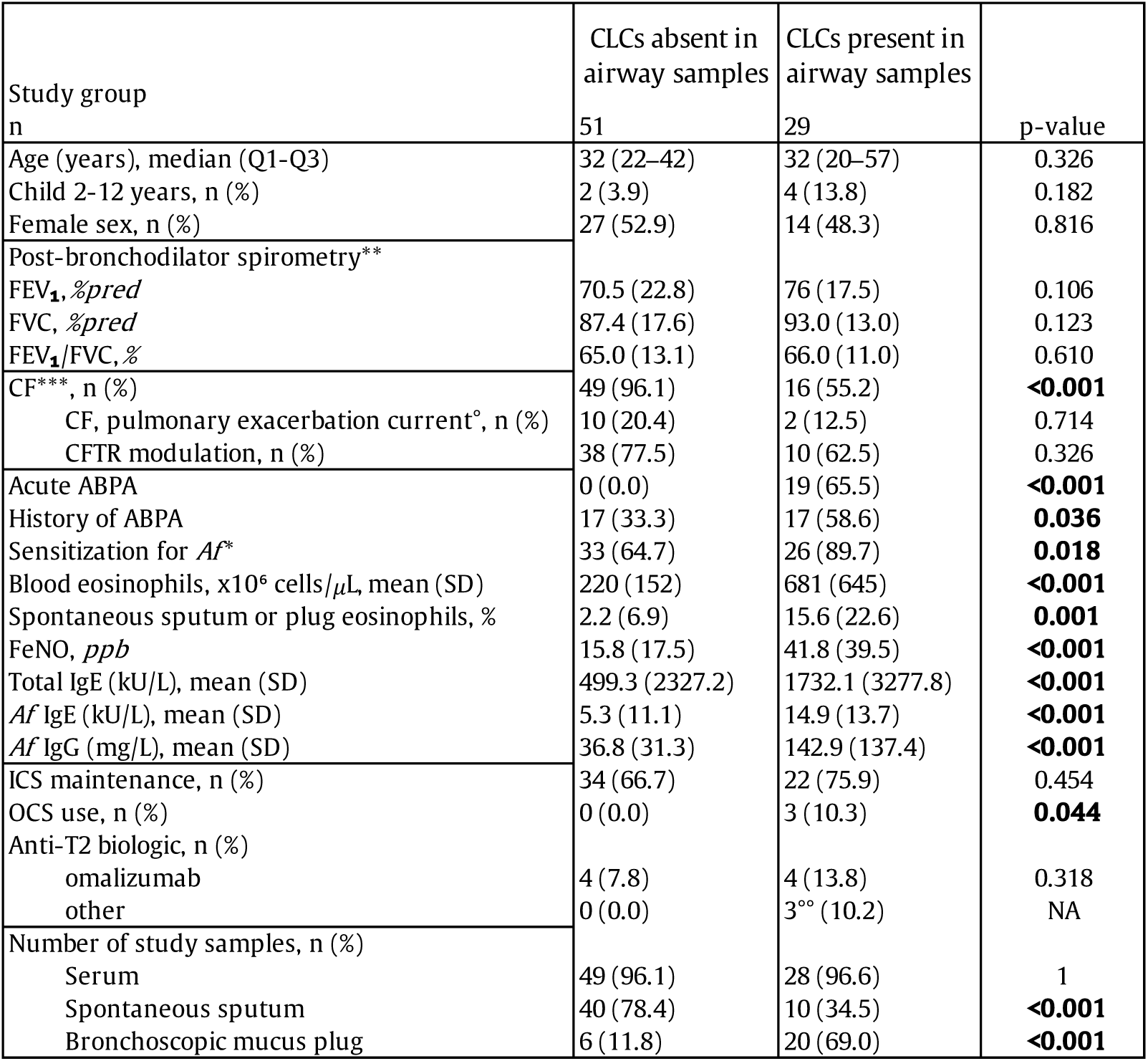
Patient characteristics stratified by Charcot-Leyden crystal (CLC) study group and substratified by underlying disease. Abbreviations: ABPA: allergic bronchopulmonary aspergillosis; Af: Aspergillus fumigatus; CF: cystic fibrosis; CFTR: cystic fibrosis transmembrane conductance regulator protein; ELX: elexacaftor; FeNO: fractional exhaled nitric oxide; FEV1 %pred: percent of predicted of forced expiratory volume in one second, FVC %pred: percent of predicted of forced vital capacity; ICS: inhaled corticosteroids; Ig: immunoglobulin; OCS: oral corticosteroids; ppb: parts per billion; WBC: white blood cell count. °Indication for oral or intravenous antibiotics. °°mepolizumab (n=1), dupilumab (n=1), tezepelumab (n=1). *Cut-off value Af IgE 0.35 kU/L. **If a post bronchodilator value was not available, the pre bronchodilator value was used. ***All subjects without cystic fibrosis have asthma. Statistical test for continuous variables: Mann-Whitney U test; for categorical variables: Fisher’s Exact test. P values < 0.05 are in bold. Note1: Each entry represents a unique clinical episode and a unique individual. Note 2: eight individuals provided a sputum and a plug sample within the same clinical episode, of whom five in ‘CLCs absent in airway samples’ and three in ‘CLCs present in airway samples’.

**SUPPLEMENT TO FIGURE 1.**
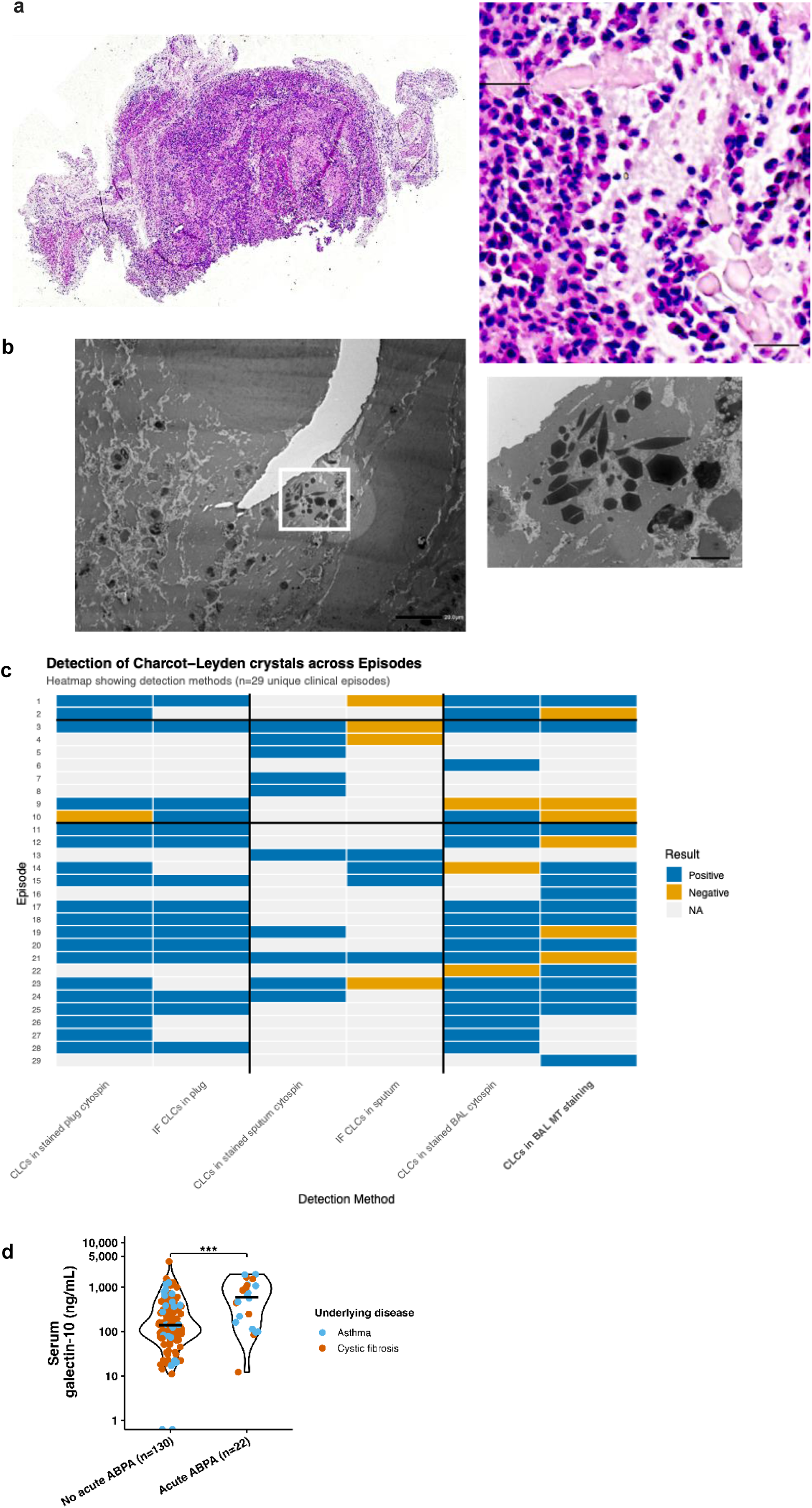
**a**, Light microscopy image of a hematoxylin and eosin staining of a mucus plug extracted from the airways of an ABPA patient. Charcot-Leyden crystals (CLCs) appear pale pink. Scale bars, 500 μm (overview image) and 20 μm (detail). **b,** CLCs embedded in a sputum plug, image acquired by transmission electron microscopy. Scale bars, 10 μm. **c,** The heatmap displays detection results for Charcot-Leyden crystals (CLCs) across 29 unique clinical episodes, organized by sample type and acute ABPA clinical status. The vertical lines separate detection methods performed on plug samples (left), spontaneous sputum samples (middle), and bronchoa lveolar lavage (BAL) samples (right). The horizont al lines separate acut e ABPA episodes (bottom) from asthma and cystic fibrosis control episodes (middle) and episodes that were excluded from ABPA comparisons, but included for CLC comparisons (top). Detection methods available in routine clinical practice are indicated in bold. Colors represent positive detection (blue), negative detection (orange), and tests not performed (light gray). BAL, bronchoalveolar lavage; CLC, Charcot –Leyden crystal; IF, immunofluorescence; MT, Masson’s trichrome. **d,** Serum Gal-10 concentration by acute ABPA diagnosis. Violin plots show Gal-10 in subjects without (n = 130, median 136 ng mL⁻¹) or with (n = 22, median 599 ng mL⁻¹) acute ABPA on a logarithmic scale. *p* = 0. 0003 ABPA, allergic bronchopulmonary aspergillosis; CLCs, Charcot-Leyden crystals; Gal-10, galectin-10. Points are colored by disease (asthma, sky blue; CF, vermilion) and shaped by sample type (spontaneous sputum, ●; plug, ×). Black crossbars indicate medians. Each sample represents one unique clinical episode. If spontaneous sputum and plug from the same episode were available (n=8), the plug s ample was selected. *p* < 0. 05, ** *p* < 0. 01, *** *p* < 0. 001, **** *p* < 0. 0001. Mann-Whitney U statistical test unless otherwise stated. See Supplementary Table 1 for full statistical details in the different

**SUPPLEMENT TO FIGURE 2.**
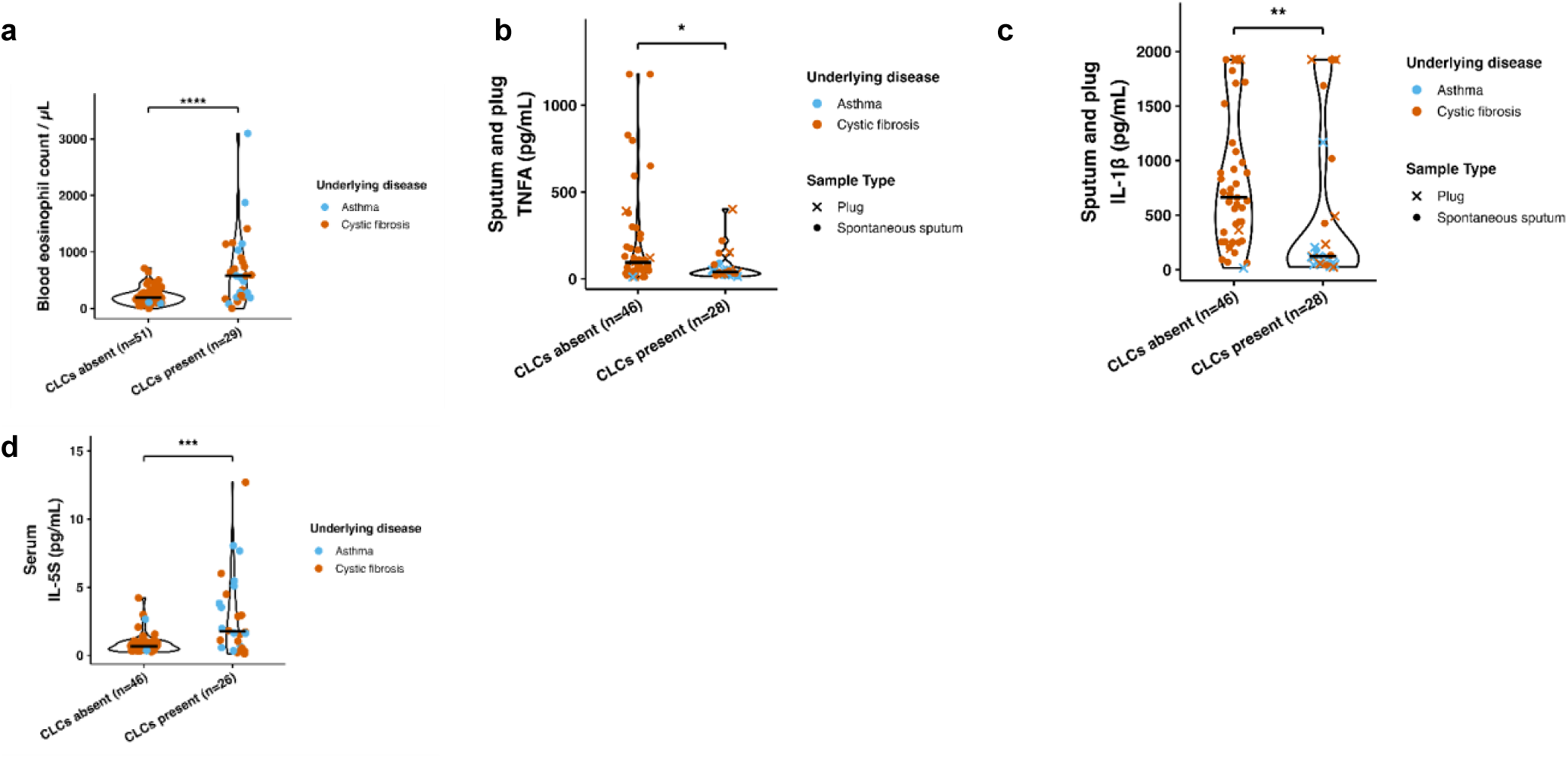
Comparison of samples with CLCs absent or present in the context of asthma and/or CF for (**a**), blood eosinophil count (median 190 vs 580/µL, *p* < 0. 0001 (**b**), TNF-α (median 95.34 vs 40.70 pg mL⁻¹, *p* = 0. 011); (**c**), IL-1ß (median 661. 65 vs 122. 55 pg mL⁻¹, *p* = 0. 001). (**d**) serum interleukin (IL)-5 (median 0.68 vs 1.76 pg mL⁻¹, *p* < 0. 001); ABPA, allergic bronchopulmonary aspergillosis; CLCs, Charcot-Leyden crystals; Gal-10, galectin-10. Ppb, parts per billion. Points are colored by disease (asthma, sky blue; CF, vermilion) and shaped by sample type (spontaneous sputum, ●; plug, ×). Black crossbars indicate medians. * *p* < 0. 05, ** *p* < 0. 01, *** *p* < 0. 001, **** *p* < 0. 0001. Mann-Whitney U statistical test unless otherwise stated.

**SUPPLEMENT TO FIGURE 3.**
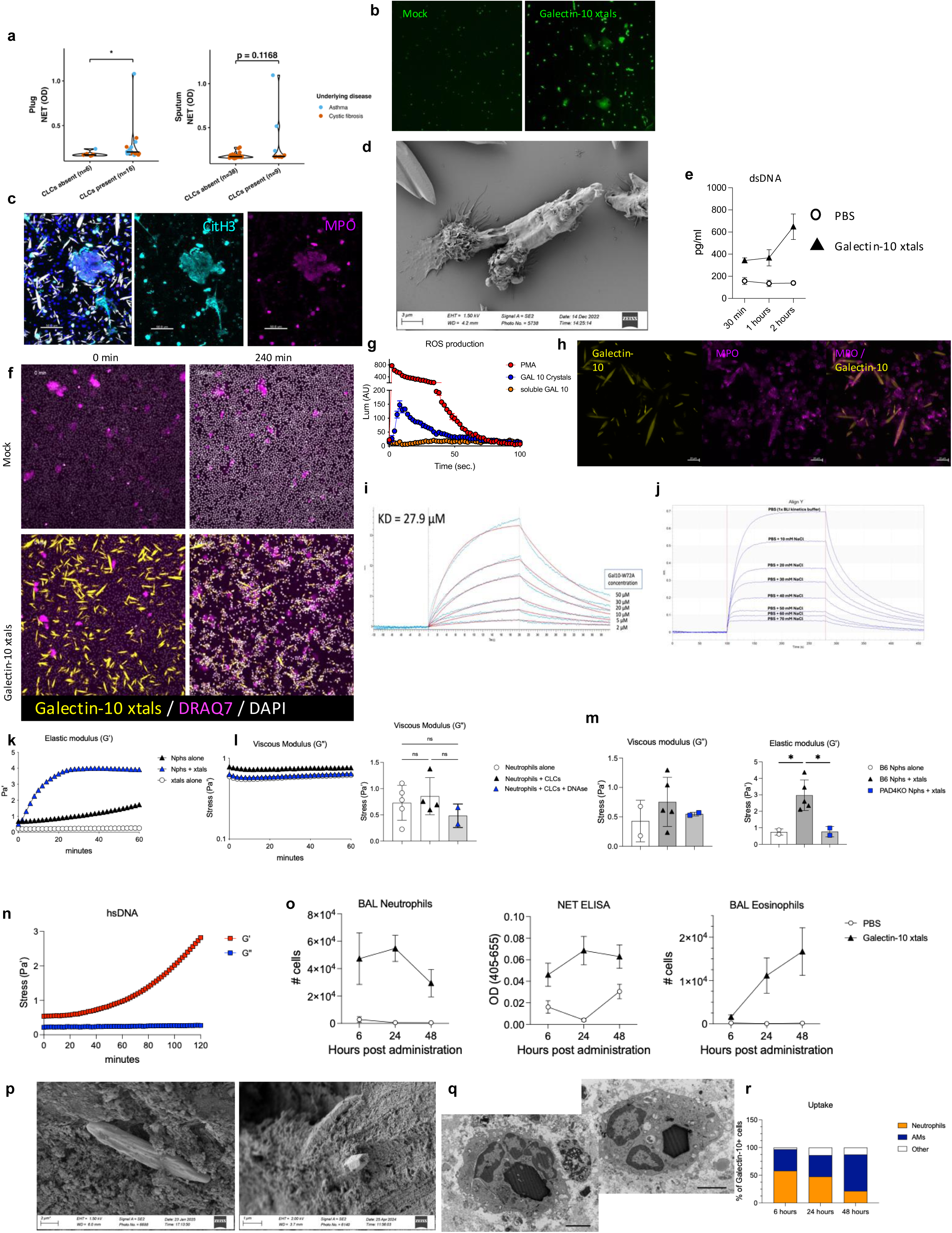
**a,** Comparison of samples with CLCs absent or present in the context of asthma and CF for NETosis via ELISA. Data was selected for plug subset (left) (median 0.17 vs 0.20, p = 0.013) and sputum subset (right) (median 0.17 vs 0.17, p = 0.117). Points are colored by disease (asthma, sky blue; CF, vermi lion) and shaped by sample type (spontaneous sputum,; plug, ×). Black crossbars indicate medians. Mann-Whitney U statistical test. **b,** Release of NETs by mouse bone-marrow derived neutrophils left unstimulated (PBS) or stimulated with 100µg/ml CLCs for 4 hours followed by staining of extracellular DNA with SYTOX Green scale bar **c,** Detection of NETs produced by mouse bone-marrow derived neutrophils stimulated with 100µg/ml CLCs for 4 hours followed by immuno-staining of Citrullinated H3 (Cit-H3), Myeloperoxidase (MPO), DAPI and detection of FITC-CLCs Scale bar, 50µm **d,** Multiple neutrophils and NETs decorate a CLC following incubation of mouse bone –marrow derived neutrophils with 100µg/ml CLCs for 4 hours, shown with scanning electron microscopy scale bar **e,** Production of dsDNA by mouse bone-marrow derived neutrophils stimulated with PBS or 100µg/ml CLCs and quantified with Quant-IT TM PicoGreen dsDNA Assay Kit (Invit rogen) **f,** Example stills of live-cell imaging at 0 hours and 4 hours (240 minutes) of DRAQ7 (a cell impermeable dye that intercalates dsDNA) in live cell imaging of human peripheral neutrophils unstimulated or stimulated with 100µg/ml CLCs for 4 hours demonstrating the clustering of neutrophils around CLCs and the abundant NET production **g,** Luminol assay demonstrates an appreciable burst of Reactive Oxygen Species (ROS) from human neutrophils stimulated with 100µg/ml CLCs **h,** Imaging of FITC-labelled Galectin-10 crystals coated with MPO after 4 hour incubation with human neutrophils. Scale bar, 20µm **i,** Interaction of dsDNA with W72A (CRD-domain mutant) Galectin-10 as demonstrated by Biolayer Interferometry (BLI) measurement **j,** Interaction of dsDNA with Galectin-10 disrupted in increasing salt concentrations as demonstrated by Biolayer Interferometry (BLI) measurement **k,** An example experiment and quantification of the elastic (G’) modulus of a hyaluoronic acid (HA) based hydrogel incubated with human neutrophils stimulated with PBS, Galectin-10 crystals, or Galectin-10 crystals alone. **l,** An example experiment and quantification of the viscous (G’’) modulus of a hyaluoronic acid (HA) based hydrogel incubated wit h human neutrophils stimulated with PBS, Galectin-10 crystals, or Galectin-10 crystals in the presence of DNAse. **m,** Quantification of the viscous (G’’) and elastic (G’) modulus of a hyaluoronic acid (HA) based hydrogel incubated with mouse neutrophils stimulated with PBS, Galectin-10 crystals. Neutrophils were isolated from WT (C57BL/6) mice or PAD4 −/− mice – deficient in the ability to make cit rullinated NETs. **n,** Detection of the viscous (G’’) and elastic (G’) modulus of a hyaluoronic acid (HA) based hydrogel incubated with herring sperm DNA (hsDNA) dem onstrating the increse in elastic, but not viscous modulus following addition of DNA. **o,** An infiltration of neutrophils, eosinophils quantified by flow cytometry and production of NETS quantified by ELISA in the br oncho-alveolar lavage fluid at 6, 24 and 48 hours following instillation of 100ug FITC-labelled CLCs (or PBS) in 70µl. **p**, Scanning electron microscopy of CLCs in a human mucus plug where porous holes in CLCs can be seen **q**, Transmission electron microscopy of a CLCs engulfed by neutrophils f rom a human mucus plug. Scale bar to add **r,** Flow cytometric quantification of uptake of FITC- labelled CLCs by neutrophils and AMs, CLC+ cells were identified by FITC signal and CLC+ neutrophils and alveolar macrophages were identified by flow cytometry and quantified in the broncho-alveolar lavage fluid at 6, 24 and 48 hours following instillation of 100ug FITC-labelled Galectin-10 crystals in 70µl.

**Supplementary figure 4.**
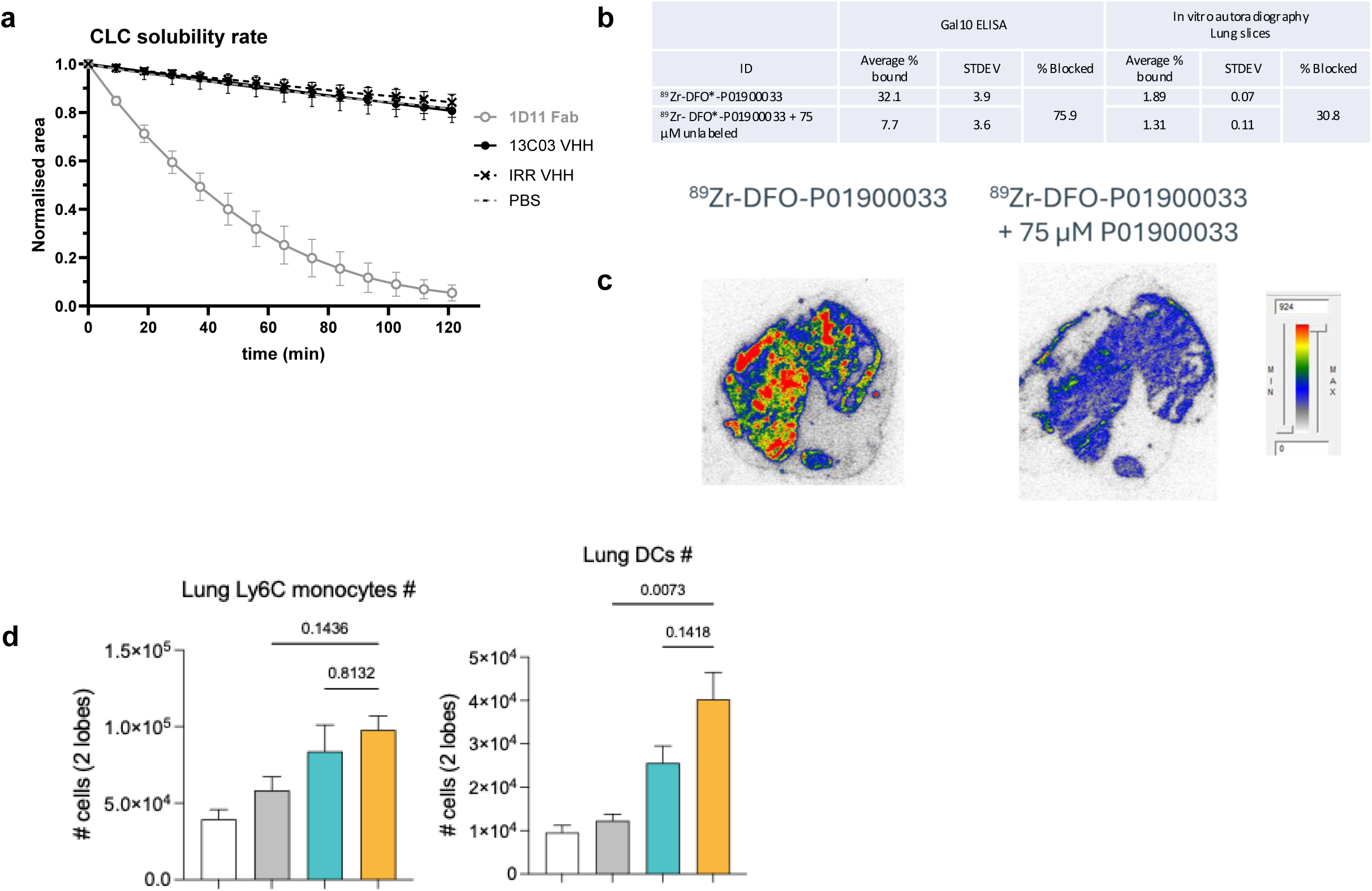
**a,** Live cell imaging of CLC dissolution by Gal10 VHHs **b,** Target binding of 89Zr-DFO*- P01900033 **c,** In vitro autoradiography 89Zr-DFO*- P01900033 **d,** Flow cytometric quantification of Ly6C + monocytes and MHC-II+ CD11c+ dendritic cells (DCs) in the lung tissue at 4 days post-administration. C57BL/6 mice were intra-tracheally administered PBS, Galectin-10 crystals, Peptide gel alone, or Peptide gel + Galectin-10 crystals.

